# COVID-19 Vaccine Rollouts and the Reproduction of Urban Spatial Inequality: Disparities Within Large U.S. Cities in March and April 2021 by Racial/Ethnic and Socioeconomic Composition

**DOI:** 10.1101/2021.11.19.21266593

**Authors:** Nicholas V. DiRago, Meiying Li, Thalia Tom, Will Schupmann, Yvonne Carrillo, Colleen M. Carey, S. Michael Gaddis

## Abstract

Rollouts of COVID-19 vaccines in the U.S. were opportunities to redress disparities that surfaced during the pandemic. Initial eligibility criteria, however, neglected geographic, racial/ethnic, and socioeconomic considerations. Marginalized populations may have faced barriers to then-scarce vaccines, reinforcing disparities. Inequalities may have subsided as eligibility expanded. Using spatial modeling, we investigate how strongly local vaccination levels were associated with socioeconomic and racial/ethnic composition as authorities first extended vaccine eligibility to all adults. We harmonize administrative, demographic, and geospatial data across postal codes in eight large U.S. cities over three weeks in Spring 2021. We find that, although vaccines were free regardless of health insurance coverage, local vaccination levels in March and April were negatively associated with poverty, enrollment in means-tested public health insurance (e.g., Medicaid), and the uninsured population. By April, vaccination levels in Black and Hispanic communities were only beginning to reach those of Asian and White communities in March. Increases in vaccination were smaller in socioeconomically disadvantaged Black and Hispanic communities than in more affluent, Asian, and White communities. Our findings suggest vaccine rollouts contributed to cumulative disadvantage. Populations that were left most vulnerable to COVID-19 benefited least from early expansions in vaccine availability in large U.S. cities.

## INTRODUCTION

### Background: COVID-19 Vaccine Rollouts in the U.S

In early 2021, state and local authorities in the United States (U.S.) vaccinated millions of people weekly against Coronavirus Disease 2019 (COVID-19).^1^ Vaccination curbed viral infection and transmission and reduced illness, hospitalization, and death from COVID-19.^2–8^ Vaccines were free countrywide regardless of health insurance coverage. Eligibility progressed in stages per state and local policy. Health care employees received first priority, followed by seniors, workers in designated occupations, and individuals with particular medical conditions.^9^

The most significant expansion in eligibility occurred during late March and April 2021. Doses remained scarce, but most jurisdictions allowed everyone age 16 and older to be vaccinated.^10–15^ On January 1, 2021, 5.5 million people had received at least one dose of a COVID-19 vaccine. That number increased to 30.3 million by February 1, 57.0 million by March 1, 111.1 million by April 1, and 153.7 million by May 1.^16^ Growth plateaued in May. Over 206.6 million U.S. residents were at least partially vaccinated by September 1; nearly half of them received their first dose in March or April. Eligibility expansion enabled rapid increases during these months.

Vaccine eligibility rules did not account for two key predictors of the burden of the COVID-19 pandemic in the U.S.: race/ethnicity and socioeconomic status (SES). At the community level, infection and mortality were more common where low-SES individuals and people of color (POC) comprised more of the population.^17–24^ At the individual level, Black and Hispanic people were disproportionately likely to experience infection, hospitalization, and death.^9, 25–34^ Socioeconomic variables partially mediated racial/ethnic disparities.

Decision-makers might have opted against conditioning vaccine eligibility on racial/ethnic or socioeconomic factors to avoid legal challenges.^35, 36^ Still, there were viable ways to use vaccine eligibility policy to mitigate inequality.^37^ For example, the Advisory Committee on Immunization Practices (ACIP) initially recommended prioritizing essential workers, which would have increased eligibility for low-SES people and POC.^38^ Authorities ultimately hewed closer to ACIP’s final recommendations, giving greater weight to advanced age.^39^

Geographic allocation may have been the most promising indirect means of addressing disparities through the rollout of COVID-19 vaccines. SES, race/ethnicity, and geography are tightly linked in the reproduction of inequality in the U.S.; socioeconomic and racial/ethnic inequalities manifest in space, usually at hyperlocal scales.^40–51^ Prioritizing local geographies in which residents had the highest risks of hardship from COVID-19 probably would have reduced mortality more than the age-based rollouts authorities chose.^52^ Regardless of whether demographic targeting was constitutional, spatial targeting could have advanced vaccine equity.

### Motivation: Early Vaccine Distribution and Cumulative Disadvantage

Authorities relied on individual initiative to distribute vaccines outside the health care workforce. This approach favored individuals with internet access, reliable transportation, and flexible schedules. States and localities used first come, first served online scheduling for scarce appointments at small numbers of sites. People with reliable internet access and white-collar jobs were better positioned to sign up. Limited locations and timed appointments were disadvantageous for people with restricted transit options and strict or uncertain work schedules, including the poor and many people with disabilities. Barriers to vaccination in March and April 2021 may have reinforced socioeconomic and racial/ethnic disparities.

Concern over vaccine hesitancy in the U.S. has abounded, but framing vaccination solely as a matter of individual choice obscures structural and material impediments. Researchers mostly attribute stagnant U.S. vaccination rates to misinformation, mistrust in institutions, and political party affiliation.^53–64^ At the same time, survey evidence suggests vaccination was linked to SES in Spring 2021.^65^ Unvaccinated respondents reported three major economic concerns: taking time off work to get the vaccine, missing work due to side effects, and out-of-pocket costs. Plausible economic determinants of vaccine uptake as eligibility first expanded suggest racial/ethnic and socioeconomic disparities may have arisen.

If disparities persisted through April 2021, vaccine rollouts contributed to cumulative disadvantage.^66, 67^ Advantages secure future advantages; inequality begets inequality—including at the neighborhood level.^43, 68–78^ Vaccine rollouts may have propelled a circular process. POC and low-SES communities were most likely to experience serious illness or die from COVID-19. Equitable vaccine distribution would have mitigated racial/ethnic and socioeconomic gaps, but early vaccine distribution did not account for these inequalities. As a result, geographic clusters of unvaccinated people could have emerged, restarting the cycle by facilitating viral transmission.^79–85^ Understanding racial/ethnic and socioeconomic vaccination disparities at the local level identifies harms that marginalized people experienced during the pandemic and helps explain the reproduction of urban spatial inequality in the U.S.^86–89^

### Overview

Our analysis provides a unique perspective on socioeconomic, racial/ethnic, and spatial disparities during the pandemic in the U.S. Numerous studies have focused on geographic dimensions of COVID-19-related inequality,^90–97^ but few have examined spatial differences in vaccination below the state level.^59, 98–100^ The temporal persistence of geographical vaccination disparities is particularly underexplored. We also contribute a novel dataset^101^ that harmonizes initially incompatible sources. And unlike many studies of COVID-19 disparities—even analyses with a geographical focus—we modeled spatial dynamics.

We tested two hypotheses. First, we hypothesized that local areas in which POC and low-SES individuals comprised more of the population had lower vaccination levels in March and April 2021. Second, we hypothesized that, despite lower starting points, the same areas had smaller increases in vaccination between March and April.

We used spatial quantitative methods to test these hypotheses. We estimated associations between vaccination levels and racial/ethnic and socioeconomic composition, adjusting for populations with early eligibility due to age or employment. We collected administrative data on vaccination by postal code, covering eight of the 10 most populous U.S. cities in March and April 2021. We combined these data with demographic estimates and geospatial data from the U.S. Census Bureau. We used spatial interpolation to reconcile reporting irregularities.

We found that, although vaccines were free regardless of health insurance coverage, local vaccination levels in March and April were negatively associated with poverty, enrollment in means-tested public health insurance (e.g., Medicaid), and the uninsured population. By April, vaccination levels in Black and Hispanic communities were only beginning to reach those of Asian and White communities in March. Increases in vaccination were smaller in socioeconomically disadvantaged Black and Hispanic communities than in more affluent, Asian, and White communities. Our findings suggest vaccine rollouts contributed to cumulative disadvantage.

## DATA AND METHODS

### Data

From online public databases maintained by state and local public health authorities, we gathered official counts of individuals with at least one dose of a COVID-19 vaccine in March and April 2021. Only geographically aggregated data were publicly available. We secured them for eight of the 10 most populous U.S. cities: New York, Chicago, Houston, Phoenix, Philadelphia, San Antonio, San Diego, and Dallas (in descending order of population). The vaccination data capture a three-week window during which eligibility expanded significantly. The number of individuals with at least one dose of a COVID-19 vaccine in the eight cities increased 34.7 percent from 4.6 million to 7.1 million during this period. We present key details of the vaccination data in Table 1; we elaborate in Section e2.1 of the online supplement.

**Table 1.** Vaccination data sources and coverage.

| City | Source | Time 1 | Time 2 |
| --- | --- | --- | --- |
| New York | New York City Department of Health and Mental Hygiene | March 22, 2021 | April 13, 2021 |
| Chicago | Chicago Department of Public Health | March 22, 2021 | April 13, 2021 |
| Houston | Texas Department of State Health Services | March 22, 2021 | April 11, 2021 |
| Phoenix | Arizona Department of Health Services | March 22, 2021 | April 13, 2021 |
| Philadelphia | Philadelphia Department of Public Health | March 21, 2021 | April 12, 2021 |
| San Antonio | Texas Department of State Health Services | March 22, 2021 | April 11, 2021 |
| San Diego | County of San Diego Health and Human Services Agency | March 21, 2021 | April 12, 2021 |
| Dallas | Texas Department of State Health Services | March 22, 2021 | April 11, 2021 |

We used two datasets from the U.S. Census Bureau. We collected demographic data from the 2015–2019 American Community Survey (ACS) Five-Year Estimates^102^ and geospatial vector data from the 2019 TIGER/Line Shapefiles.^103^ We provide further detail on these sources in Sections e2.2 and e2.3 of the online supplement.

### Unit of Analysis

For brevity and interpretability, we refer to our units of analysis as ZIP Codes, the name for postal codes in the U.S. The units of analysis were based on ZIP Codes, but reporting irregularities made ZIP Codes themselves inviable. Where necessary, we used overlay interpolation^104, 105^ to exclude populations residing outside city limits. We provide extensive detail on the units of analysis and interpolation in Section e3 of the online supplement.

### Independent Variables

#### Vaccination Priority Populations

We accounted for vaccination priority regulations by adjusting for populations of health care workers and seniors. Specific estimates were unavailable for health care workers, but ACS provided estimated counts of individuals employed in “health care and social assistance.” We also adjusted for the share of the population age 65 or older. These variables were the best available measures of the first groups prioritized for vaccination. We include more information on these variables in Section e2.2 of the online supplement.

#### Socioeconomic Composition

To examine the dependent variable’s association with socioeconomic composition, we included four indicators of SES. Two independent variables estimated health insurance status. Health insurance coverage was not universal in the U.S. as of the COVID-19 pandemic, and medical care remained expensive and stratified compared to other rich countries.^106, 107^ We included variables estimating the share of the population enrolled in Medicaid or other means-tested public health insurance and the share without health insurance altogether. Together, these variables captured populations that were among the least integrated into the U.S. health care system. We also included variables estimating the shares of the population under the federal poverty line and without internet access. We included the latter because making appointments online was usually the best way to secure a vaccine in early 2021. We include more information on our SES variables in Section e2.2 of the online supplement.

#### Racial/Ethnic Composition

We accounted for racial/ethnic composition because racism causes health inequity in the U.S.^108–115^ Although race/ethnicity itself cannot cause anything, distributive systems that allocate resources according to racial/ethnic hierarchies create disparities among racial/ethnic groups.^116–120^ These disparities often surface net of SES. Including measures of racial/ethnic composition in our models enabled us to examine its direct association with vaccination, adjusting for SES.

Racism, however, is more than a conditional association between an outcome and racial/ethnic composition.^121–126^ It undergirds the gamut of U.S. social, economic, and political processes. The distributions of socioeconomic covariates and unobserved mechanisms were racialized. We analyzed racism in the aggregate by considering direct and indirect pathways— mainly through simulations, described below and in Section e4.3 of the online supplement.

From ACS racial/ethnic categories, we created variables measuring the estimated populations of four mutually exclusive, non-exhaustive racial/ethnic groups: Asian, Black, Hispanic, and White. We defined Hispanic as Hispanic, Latino, or Spanish origin, of any race(s). We defined Black, Asian, and White as non-Hispanic and Black or African American alone, Asian alone, and White alone, respectively. This approach implies a fifth category comprised of non-Hispanic individuals of multiple races or any other race alone. The racial/ethnic variables did not sum to one (100 percent) unless the estimated population of the fifth category was zero.

We include more information on our framework for race/ethnicity and racial/ethnic variables in Section e2.2 of the online supplement.

### Dependent Variable

The dependent variable approximated the share of each ZIP Code’s vaccine-eligible population that was partially or fully vaccinated against COVID-19. We calculated it by dividing the estimated number of residents with at least one dose of an approved vaccine by the estimated population age 15 and older. This denominator was the best available measure of the population to whom agencies were authorized to administer vaccines in March and April 2021. More information on the dependent variable is available in Section e2 of the online supplement.

### Spatial-Statistical Analysis

We estimated population-weighted regressions^127^ with conventional adjustments for spatial clustering. We report spatial error models (SEMs) estimated by maximum likelihood.^128–133^ Standard linear models (SLMs) are ill-suited to estimate associations that vary across space. In this analysis, spatial heterogeneity could have arisen from unmeasurable factors such as COVID-19 exposure, hyperlocal idiosyncrasies in the effects or implementation of vaccination policies, and cultural influences. Standard tests^134, 135^ strongly suggested SLMs exhibited spatial heterogeneity in our setting. We estimated SEMs with row-standardized *k* nearest-neighbor weights (*k* = 8).^136–138^ As the Moran’s *I* test statistics in Table 3 demonstrate, the SEMs eliminated the residual spatial clustering that emerged in the SLMs. The models incorporated city fixed effects to adjust for unmeasured variables that were constant among ZIP Codes within each city,^139^ including elements of vaccination policies. Because multiple cities were in Texas, we calculated heteroskedasticity-robust standard errors clustered by state.^140^

To illustrate the estimated associations, we simulated outcomes at representative values in the racial/ethnic and socioeconomic distributions of the sample. This approach resembled a marginal effects analysis but accounted for spatial clustering and yielded an overall average rather than a unit-level estimate.^141–144^ We present eight simulated scenarios: ZIP Codes with high Black populations and (1) low SES or (2) high SES; high Hispanic populations and (3) low SES or (4) high SES; high Asian populations and (5) low SES or (6) high SES; and high White populations and (7) low SES or (8) high SES. We defined low and high levels as below the 10th and above the 90th within-city percentiles, respectively.

We provide additional details on all aspects of our analytical approach, including the models and simulations, in Section e4 of the online supplement.

## RESULTS

### Descriptive Findings

In Table 2, we present descriptive statistics at the ZIP Code level. On average across all 552 ZIP Codes, 28.0 percent of the population in March and 42.4 percent of the population in April had at least one dose of a COVID-19 vaccine, with a mean difference of 14.5 percentage points between March and April. Other than Philadelphia and San Diego, each city’s mean vaccination level fell within a two-point range (27–29 percent) in March and a five-point range (40–45 percent) in April. Although there was some variation between cities, vaccination levels varied considerably more across ZIP Codes within cities (see Figure 1). In March, the standard deviation in vaccination levels was 3.0 percent between cities and 8.8 percent within cities; in April, it was 4.2 percent between cities and 11.9 percent within cities. The mean difference between the 10th and 90th percentiles of vaccination levels across cities was 21.6 percent in March and 31.0 percent in April.

**Figure 1.**
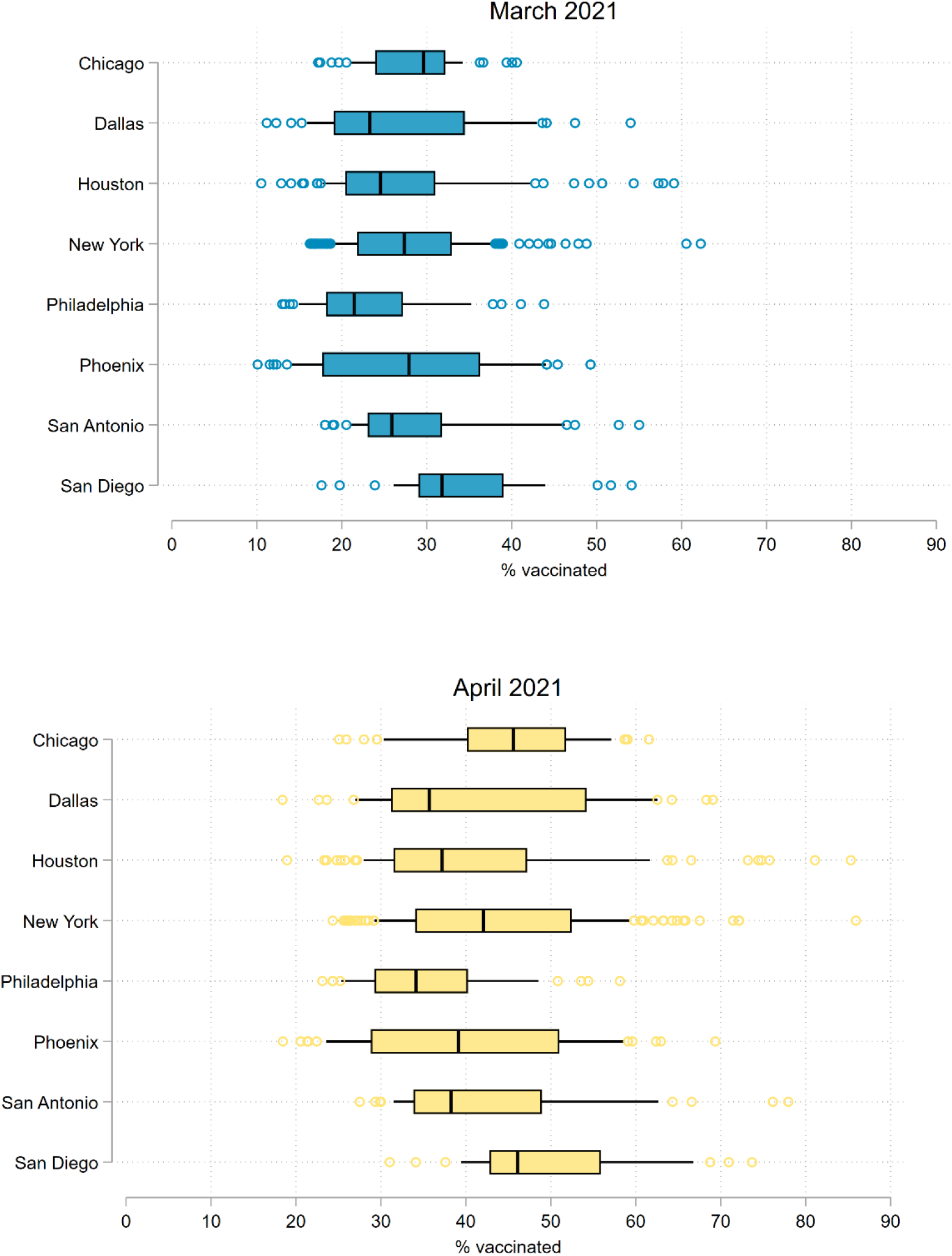
COVID-19 vaccination levels in the population age 15 and older of ZIP Codes in eight large U.S. cities, March and April 2021. Note: Figures are box-and-whisker plots of vaccination levels in *N* = 552 ZIP Codes across eight of the 10 most populous U.S. cities. The boxes represent interquartile ranges. The vertical lines represent medians. The horizontal lines extend from the 10th to the 90th percentiles. Circles represent observations below the 10th and above the 90th percentiles. The “% vaccinated” is the percent of the population age 15 and older with at least one dose of a COVID-19 vaccine.

**Table 2.** Descriptive statistics on COVID-19 vaccination and population composition in ZIP Codes within and across eight large U.S. cities, March and April 2021.

|  | *M* | *SD* |  | *M* | *SD* |  | *M* | *SD* |
| --- | --- | --- | --- | --- | --- | --- | --- | --- |
| **New York (*n* = 175)** |  |  | **Chicago (*n* = 53)** |  |  | **Houston (*n* = 99)** |  |  |
| % vaccinated, March | 28.18 | 7.94 | % vaccinated, March | 28.70 | 5.50 | % vaccinated, March | 27.23 | 10.03 |
| % vaccinated, April | 43.60 | 11.71 | % vaccinated, April | 45.08 | 9.46 | % vaccinated, April | 40.97 | 13.64 |
| % vaccinated, difference | 15.42 | 5.24 | % vaccinated, difference | 16.38 | 5.21 | % vaccinated, difference | 13.75 | 4.03 |
| % 65+ | 14.98 | 5.09 | % 65+ | 12.55 | 4.06 | % 65+ | 10.32 | 3.09 |
| % health care workers | 17.42 | 6.49 | % health care workers | 13.93 | 3.73 | % health care workers | 10.81 | 3.31 |
| % under poverty line | 15.91 | 9.42 | % under poverty line | 17.93 | 9.90 | % under poverty line | 18.33 | 9.36 |
| % w/ Medicaid, etc. | 16.61 | 9.96 | % w/ Medicaid, etc. | 12.98 | 9.48 | % w/ Medicaid, etc. | 6.35 | 4.13 |
| % w/o health insurance | 8.16 | 4.40 | % w/o health insurance | 10.13 | 5.97 | % w/o health insurance | 25.09 | 12.31 |
| % w/o internet access | 14.62 | 6.47 | % w/o internet access | 16.34 | 9.10 | % w/o internet access | 16.51 | 10.85 |
| % Black | 19.82 | 23.39 | % Black | 29.67 | 33.53 | % Black | 22.73 | 18.63 |
| % Hispanic | 26.37 | 19.34 | % Hispanic | 22.43 | 21.94 | % Hispanic | 42.58 | 22.27 |
| % Asian | 14.77 | 13.96 | % Asian | 7.87 | 8.61 | % Asian | 6.56 | 6.23 |
| **Phoenix (*n* = 50)** |  |  | **Philadelphia (*n* = 46)** |  |  | **San Antonio (*n* = 48)** |  |  |
| % vaccinated, March | 27.78 | 11.00 | % vaccinated, March | 23.30 | 7.65 | % vaccinated, March | 29.10 | 9.04 |
| % vaccinated, April | 40.17 | 13.48 | % vaccinated, April | 35.58 | 8.74 | % vaccinated, April | 42.21 | 11.90 |
| % vaccinated, difference | 12.38 | 3.19 | % vaccinated, difference | 12.28 | 2.01 | % vaccinated, difference | 13.11 | 3.33 |
| % 65+ | 11.32 | 4.35 | % 65+ | 14.18 | 4.61 | % 65+ | 11.90 | 3.24 |
| % health care workers | 12.08 | 2.16 | % health care workers | 20.63 | 4.38 | % health care workers | 14.06 | 2.31 |
| % under poverty line | 16.82 | 10.04 | % under poverty line | 22.25 | 11.17 | % under poverty line | 16.55 | 8.85 |
| % w/ Medicaid, etc. | 12.38 | 7.14 | % w/ Medicaid, etc. | 15.52 | 9.37 | % w/ Medicaid, etc. | 4.99 | 3.01 |
| % w/o health insurance | 14.37 | 8.32 | % w/o health insurance | 8.74 | 3.58 | % w/o health insurance | 18.89 | 8.42 |
| % w/o internet access | 13.49 | 9.36 | % w/o internet access | 18.16 | 8.49 | % w/o internet access | 15.69 | 9.88 |
| % Black | 6.04 | 4.29 | % Black | 38.51 | 30.97 | % Black | 7.13 | 7.67 |
| % Hispanic | 37.30 | 23.95 | % Hispanic | 11.99 | 13.48 | % Hispanic | 61.79 | 20.90 |
| % Asian | 3.96 | 2.77 | % Asian | 6.96 | 5.82 | % Asian | 2.85 | 2.72 |
| **San Diego (*n* = 33)** |  |  | **Dallas (*n* = 48)** |  |  | **Overall (*n* = 552)** |  |  |
| % vaccinated, March | 34.16 | 8.45 | % vaccinated, March | 27.02 | 10.40 | % vaccinated, March | 27.95 | 9.00 |
| % vaccinated, April | 50.30 | 10.81 | % vaccinated, April | 42.04 | 13.92 | % vaccinated, April | 42.44 | 12.34 |
| % vaccinated, difference | 16.13 | 3.45 | % vaccinated, difference | 15.02 | 4.72 | % vaccinated, difference | 14.48 | 4.56 |
| % 65+ | 13.18 | 4.36 | % 65+ | 10.55 | 5.72 | % 65+ | 12.75 | 4.81 |
| % health care workers | 12.87 | 2.00 | % health care workers | 11.22 | 2.42 | % health care workers | 14.58 | 5.45 |
| % under poverty line | 11.98 | 6.68 | % under poverty line | 17.11 | 9.02 | % under poverty line | 17.08 | 9.64 |
| % w/ Medicaid, etc. | 9.63 | 7.17 | % w/ Medicaid, etc. | 5.11 | 3.89 | % w/ Medicaid, etc. | 11.52 | 9.04 |
| % w/o health insurance | 8.11 | 5.43 | % w/o health insurance | 24.26 | 11.89 | % w/o health insurance | 14.33 | 10.65 |
| % w/o internet access | 7.03 | 5.30 | % w/o internet access | 18.29 | 12.72 | % w/o internet access | 15.28 | 9.26 |
| % Black | 5.34 | 4.40 | % Black | 22.88 | 19.50 | % Black | 19.89 | 23.32 |
| % Hispanic | 26.84 | 21.38 | % Hispanic | 36.52 | 20.75 | % Hispanic | 32.68 | 23.95 |
| % Asian | 15.39 | 11.40 | % Asian | 5.23 | 9.70 | % Asian | 9.18 | 10.79 |

**Note:** *N* = 552 ZIP Codes across eight of the 10 most populous U.S. cities. “Health care workers” refers to individuals employed in health care and social assistance. “Medicaid, etc.” refers to Medicaid or any other means-tested public health insurance. The “% vaccinated” is the percent of the population age 15 and older with at least one dose of a COVID-19 vaccine.

### Model Estimates

In Table 3, we summarize the results of the SEMs with all independent variables for three outcomes: March vaccination levels, April vaccination levels, and the difference between March and April vaccination levels. In both March and April, four variables were significantly associated with the dependent variable. The first, the percent of the population age 65 and older, reflects the policy choice to place older individuals among the earliest priority groups. The other three variables were measures of socioeconomic composition: the shares of the population under the poverty line, with means-tested public health insurance, and without health insurance. Adjusting for vaccination priority populations and racial/ethnic composition, markers of low SES were negatively associated with vaccination levels. In April, vaccination levels were positively associated with the Asian share of the population.

**Table 3.** Spatial error model (SEM) estimates of COVID-19 vaccination levels in the population age 15 and older of ZIP Codes across eight large U.S. cities, March and April 2021.

|  | (1)<br>% vaccinated, March | (2)<br>% vaccinated, April | (3)<br>Difference |
| --- | --- | --- | --- |
| **Vaccination priority populations** |  |  |  |
| % 65+ | 0.593***<br>(0.048) | 0.470***<br>(0.075) | -0.122*<br>(0.054) |
| % health care workers | 0.147<br>(0.257) | -0.063<br>(0.309) | -0.201***<br>(0.055) |
| **Socioeconomic composition** |  |  |  |
| % under poverty line | -0.102*<br>(0.051) | -0.138**<br>(0.051) | -0.039<br>(0.023) |
| % w/ Medicaid, etc. | -0.102***<br>(0.024) | -0.127**<br>(0.046) | -0.021<br>(0.029) |
| % w/o health insurance | -0.418***<br>(0.039) | -0.655***<br>(0.053) | -0.234***<br>(0.023) |
| % w/o internet access | -0.040<br>(0.051) | -0.036<br>(0.060) | 0.003<br>(0.011) |
| **Racial/ethnic composition** |  |  |  |
| % Black | -0.111<br>(0.061) | -0.132<br>(0.084) | -0.021<br>(0.025) |
| % Hispanic | 0.041<br>(0.035) | 0.076<br>(0.041) | 0.036***<br>(0.010) |
| % Asian | 0.101<br>(0.067) | 0.230*<br>(0.103) | 0.127***<br>(0.037) |
| **Residual Moran's I** |  |  |  |
| Standard linear model (SLM) | 0.250*** | 0.222*** | 0.202*** |
| Spatial error model (SEM) | 0.027 | 0.014 | -0.015 |

**Note:** SEMs estimated by maximum likelihood with row-standardized nearest-neighbor spatial weighting ( $k = 8$ ). $N = 552$ ZIP Codes across eight of the 10 most populous U.S. cities. City fixed effects (reference: New York) and constant terms not shown. Percentages scaled from zero to one. All models weighted by estimated population age 15 and older. Heteroskedasticity-robust standard errors clustered by state in parentheses. \*\*\* $p < 0.001$ ; \*\* $p < 0.01$ ; \* $p < 0.05$ . Moran's $I$ $p$ -values calculated by permutation bootstrap (9,999 iterations). "Health care workers" refers to individuals employed in health care and social assistance. "Medicaid, etc." refers to Medicaid or any other means-tested public health insurance. The "% vaccinated" is the percent of the population age 15 and older with at least one dose of a COVID-19 vaccine.

Five variables were significantly associated with differences in vaccination between March and April. The shares of the population age 65 and older and employed in health care were associated with smaller increases. These associations probably reflect that these prioritized populations were widely vaccinated by the end of March. The Hispanic and Asian population shares were associated with larger increases in vaccination levels. The share of the population without health insurance was associated with smaller increases in vaccination levels.

As we detail in Tables e4.1 and e4.2 in the online supplement, we examined associations step-wise for socioeconomic and racial/ethnic composition. Racial/ethnic composition measures were often statistically significant in the absence of covariates measuring socioeconomic composition. When we included socioeconomic variables, however, the coefficients of the racial/ethnic variables were indistinguishable from zero. We further discuss implications below and in Section e4.3 of the online supplement.

### Simulated Outcomes

The simulations, illustrated in Figure 2 and in Figure e4.2 in the online supplement, contextualize relationships between racial/ethnic and socioeconomic composition. At both time points regardless of racial/ethnic composition, vaccination levels were higher where SES was higher. Socioeconomic disparities in vaccination were smaller where there was a high White population and larger where there were high Black, Hispanic, or Asian populations. In March, the highest vaccination levels (36.1 percent) were associated with high White populations and high SES; the lowest levels (17.7 percent) were associated with high Black populations and low SES. In April, the highest vaccination levels (53.8 percent) were associated with high Asian populations and high SES; the lowest levels (27.5 percent) were associated with high Black populations and low SES.

**Figure 2.**
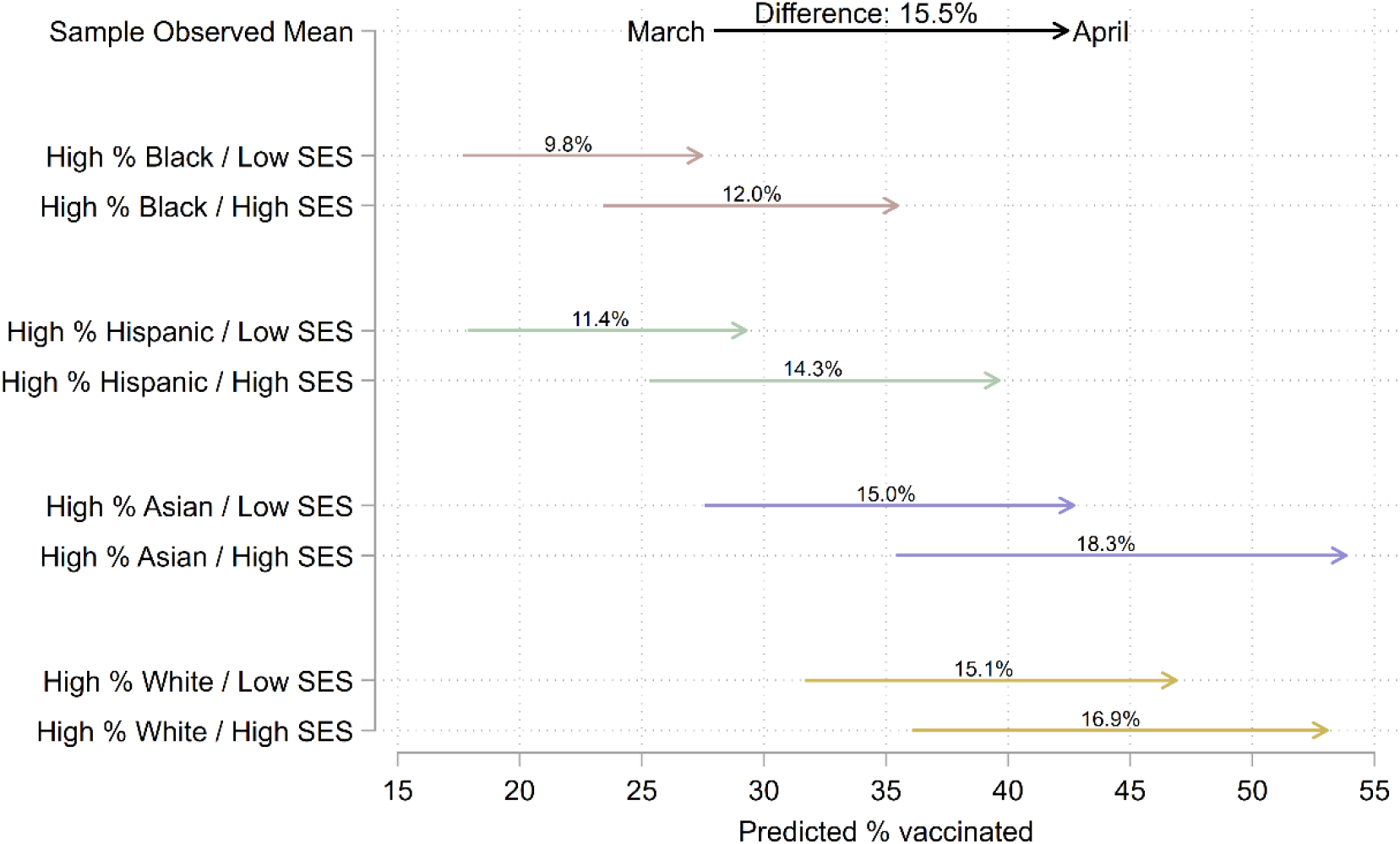
Simulated COVID-19 vaccination levels by racial/ethnic and socioeconomic composition in the population age 15 and older of ZIP Codes across eight large U.S. cities, March and April 2021. **Note**: This figure illustrates simulated sample-wide means assuming each ZIP Code had a given socioeconomic and racial/ethnic composition. We defined low and high levels as below the 10th and above the 90th within-city percentiles, respectively. We defined SES levels by setting all four SES variables to the same within-city percentiles within each scenario. We set other independent variables to within-city averages in each scenario. We include the true (observed) sample-wide average values of the dependent variable on the top row for comparison. The “% vaccinated” is the percent of the population age 15 and older with at least one dose of a COVID-19 vaccine.

Across racial compositions, the simulated change in vaccination levels between March and April was larger in high-SES than low-SES ZIP Codes, as indicated by the numbers above each line in Figure 2. Vaccination levels increased most (18.3 percentage points) where there were high Asian populations with high SES, followed by high White populations with high SES (16.9 percentage points). Vaccination levels increased least (9.8 percentage points) where there were high Black populations with low SES, followed by high Hispanic populations with low SES (11.4 percentage points).

## DISCUSSION

### Key Findings

We examined COVID-19 vaccination in eight of the 10 most populous cities in the U.S. In March and April 2021, vaccination levels varied more within cities—across ZIP Codes—than between cities. This finding suggests differences in state and local eligibility criteria contributed negligibly to disparities. Our models and simulations confirmed our hypotheses that ZIP Codes with higher shares of POC and low-SES individuals had lower vaccination levels and smaller increases over time. We now turn to three key findings.

Our finding that measures of racial/ethnic composition were statistically insignificant in the presence of socioeconomic covariates does not rule out racial/ethnic disparities. It suggests economic inequality and access to health insurance were fundamental mechanisms of local racial/ethnic gaps in vaccination. Furthermore, the relative magnitudes of the racial/ethnic variables’ coefficients were sometimes nearly as large as those of SES variables, albeit with slightly larger standard errors. Given the distribution of SES, ZIP Codes with high Black or Hispanic populations were associated with lower vaccination levels than those with high Asian or White populations.

Unlike internet access, measures of health insurance coverage were consistently associated with lower vaccination outcomes. This finding is surprising because internet access but not health insurance was directly tied to obtaining vaccine appointments. The insurance-related variables may capture multiple unobserved mechanisms: unfamiliarity with the medical system, perhaps due to reduced or discriminatory encounters with providers and insurers; incomplete or inaccurate information, including unawareness or skepticism that vaccines were free; and employment or other economic circumstances that impeded getting vaccinated or recovering from side effects. Survey or interview data may clarify individual-level mechanisms. Nonetheless, our results show that residents of large U.S. cities who had tenuous connections to the health care system were less likely to benefit from an intervention that was free to all regardless of insurance coverage.

While several inequalities increased from March to April, one waned. ZIP Codes with high Hispanic populations were associated with larger increases in vaccinations, adjusting for other demographic and socioeconomic factors. Still, accounting for socioeconomic distributions, Hispanic communities were left behind overall as vaccination eligibility expanded.

### Limitations

This study has several limitations. Authorities published vaccination data by ZIP Code only. Because ZIP Codes are suboptimal units for measuring inequality, disparities may be understated in this analysis. Representing ZIP Codes as areal polygons is distortive, potentially leading to measurement error.^145–152^ Furthermore, while they afford more local vantage points than states and counties, ZIP Codes cannot reveal finer, neighborhood-level dynamics. Our units of analysis averaged 38,123 residents, and one-quarter exceeded 50,000. At this scale, observations had substantial within-unit variation and relatively low between-unit variation, likely obscuring disparities.^52, 86, 89, 153–158^ We further discuss the analytical limitations of ZIP Codes in Section e3.1 of the online supplement.

The absence of individual-level data limited this analysis, but geographically aggregated data also presented advantages. It is difficult to determine how much our results reflected differential vaccine eligibility across ZIP Codes. We adjusted for key prioritized populations, however, and by mid-April eligibility was approaching universal among U.S. adults. In addition, the complete administrative data we used was more comprehensive than small surveys of self-reported behavior. Spatial analysis could also be optimal for guiding policy. Allocating resources geographically may be less resource-intensive than focusing on demographic subgroups. And, as we highlight above, spatial targeting is an effective tool for health equity.

## CONCLUSION

Even as the number of vaccinated individuals increased by 7.1 million (34.7 percent) in the large U.S. cities we studied, COVID-19 vaccination lagged in marginalized communities from late March through mid-April 2021. Vaccination gaps increased between low-and high-SES communities and between White or Asian and Black or Hispanic communities. The spatial clustering of unvaccinated individuals probably led to further public health issues.

Our findings suggest vaccination rollouts contributed to cumulative disadvantage at the community—and likely individual—level. Populations that experienced the highest burdens of infection and mortality from COVID-19 before vaccines were available had lower levels of vaccination during restricted vaccine eligibility. Gaps persisted or widened as eligibility first expanded. These disparities may have contributed to a bifurcated recovery in which advantaged communities began to move on from the COVID-19 pandemic while marginalized people continued to suffer.

## Data Availability

All data produced in the present study are available upon reasonable request to the authors.

## Online supplement

### e1 Introduction

In this supplement to the main text of our study, “COVID-19 vaccine rollouts and the reproduction of urban spatial inequality: disparities within large U.S. cities in March and April 2021 by racial/ethnic and socioeconomic composition,” we clarify our approach and contributions by detailing our data and methods. We used spatial quantitative methods to analyze a novel data set that harmonized previously incompatible administrative, demographic, and geospatial data. These data and methods enabled us to test for socioeconomic and racial/ethnic disparities in COVID-19 vaccination across jurisdictions and reporting agencies.

The main purpose of this supplement is to elaborate on the rationale and decisions that led to the final study. In addition to this document, interested readers can access online code and data to replicate our analysis.^1^ We gathered, wrangled, interpolated, and analyzed data using version 4.0.4 of the R statistical programming language and software environment,^2^ making extensive use of the Tidyverse.^3^ Throughout this supplement, we cite other R packages that we used. We created Figures e3.1 and e4.1 in R^4^ and all other figures using version 16.1 of Stata/MP software.^5^

Another purpose of this supplement is to argue that, in addition to the study’s empirical findings, our data and methods are substantial contributions. Because of inconsistent reporting, considerable analysis and computation were necessary to compile a high-quality data set measuring local vaccination outcomes across jurisdictions. Once we created the data set, the need to model spatial patterns became evident. Administrative obstacles and empirical dynamics expanded the study’s scope beyond the routine boundaries of observational quantitative research.

Even with appropriate adjustments, however, the absence of true neighborhood-level or other hyperlocal data from public sources limited our analysis. Conducted at the ZIP Code level, the study is the best feasible alternative to our original aim of analyzing disparities across jurisdictions by neighborhood—rather than by states or counties, which predominated in media coverage of vaccination rates at the time we began the analysis. Substantial, unnecessary barriers to measuring local inequality in COVID-19 vaccination were in place during the pandemic. Policymakers and administrators should work to reduce them. Reporting data across agencies at the same hyperlocal scale would better support analysis and resource allocation as long as vaccination disparities persist. It would also improve the reporting infrastructure for public health and other sociologically pertinent data.

The rest of this supplement is divided into four sections. In Section e2, we detail the sources, coverage, and definitions of the raw data we collected. We also specify the constructs we sought to measure, define the variables we created accordingly, and assess validity and limitations. In Section e3, we discuss the spatial scale of the study and resulting analytical obstacles and limitations, including measurement error, and detail the interpolation procedures we used to harmonize incompatible raw data. In Section e4, we explain our modeling and estimation strategy, including how we detected and accommodated spatial relationships, and explain the simulation-based approach we used to facilitate interpretation.

**Table e2.1:** Vaccination data sources and coverage (expanded)

| City | Source | As of (year 2021) |  | Universe |
| --- | --- | --- | --- | --- |
| New York | New York City Department of Health and Mental Hygiene | 22 March | 13 April | New York City residents of MODZCTA |
| Chicago | Chicago Department of Public Health | 22 March | 13 April | Chicago residents of ZIP Code |
| Houston | Texas Department of State Health Services | 22 March | 11 April | Texas residents of ZIP Code |
| Phoenix | Arizona Department of Health Services | 22 March | 13 April | Arizona residents of ZIP Code |
| Philadelphia | Philadelphia Department of Public Health | 21 March | 12 April | Philadelphia residents of ZIP Code |
| San Antonio | Texas Department of State Health Services | 22 March | 11 April | Texas residents of ZIP Code |
| San Diego | County of San Diego Health and Human Services Agency | 21 March | 12 April | San Diego County residents of ZIP Code |
| Dallas | Texas Department of State Health Services | 22 March | 11 April | Texas residents of ZIP Code |

### e2 Data

#### e2.1 Administrative records

From online public databases maintained by state and local public health authorities, we gathered official counts of individuals with at least one dose of a COVID-19 vaccine in March and April 2021. We secured these data for eight of the 10 most populous U.S. cities: New York, Chicago, Houston, Phoenix, Philadelphia, San Antonio, San Diego, and Dallas (in descending order of population). Suitable data were unavailable for Los Angeles and San José, the second and 10th most populous cities, respectively. We summarize the sources and other details of the vaccination data in Table e2.1.^e^

Only geographically aggregated data were publicly available. For each city except New York, agencies reported vaccination counts aggregated by ZIP Codes (U.S. postal codes) of residence, or by related units known as ZIP Code Tabulation Areas (ZCTAs).^f^ For New York, data were aggregated by Modified ZIP Code Tabulation Areas (MODZCTAs), proprietary spatial units that merge less populous ZCTAs with adjacent ZCTAs “to allow more stable estimates of population size for rate calculation.”^6^ Using an official crosswalk file,^7^ we harmonized data released by ZIP Code or ZCTA with the MODZCTAs.

#### e2.2 Demographic surveys

The 2015–2019 American Community Survey (ACS) Five-Year Estimates were the source of all demographic data in the analysis. Fielded annually by the U.S. Census Bureau (USCB), the ACS provides current, reliable, and representative estimates of population characteristics at various geographic scales.^8^ In preparation for the spatial processing detailed in Section e3.2, we collected all ACS tables listed in Table e2.2 by census block group (CBG).^g^ We also collected table B01001 by city and by ZCTA.^h^ We used the USCB application programming interface (API) to gather the ACS estimates.^9^

We detail the demographic variables we calculated from the ACS Table e2.2.^i^ In addition to the denominator for the outcome variables and the population weights, we used ACS data to compute all independent variables, which measured vaccine priority populations, socioeconomic composition, and racial/ethnic composition.

##### Socioeconomic status (SES) and health care

We conceptualized socioeconomic status (SES) through the conventional sociological lens of life chances,^10, 11^ or individuals’ likelihood of “gaining access to scarce and valued outcomes.”^12(p32)^ In this analysis, the scarce and valued outcome—COVID-19 vaccination—was health-related and facilitated by internet access. We adjusted for corresponding SES variables, in addition to poverty levels and vaccine priority populations.

Several independent variables measured relationships to the health care system. We partially accounted for the effects of vaccination priority regulations by adjusting for the population of health care workers. Population estimates were unavailable for this exact group, but ACS data allowed us to closely approximate them. We adjusted for the percent of the civilian employed population age 16 or older that worked in “health care and social assistance.”^13^ This category included employees of hospitals, medical practices, chiropractic practices, dental practices, optometry practices, outpatient and home health care services, nursing and residential care facilities, and other health settings. It also included employees of social service providers, child care services, and other “social assistance” professions. These social assistance workers were typically excluded from early priority groups for vaccination. The population employed in “health care and social assistance” industries was the best available proxy for health care workers, but our variable is effectively an estimate with error.

We also included two independent variables measuring health insurance status. The first was the percent of the population enrolled in Medicaid or other means-tested public health insurance. This variable comprised individuals who had “Medicaid, Medical Assistance, or any kind of government-assistance plan for those with low incomes or a disability.”^14p73^ It included individuals who had one of these types of insurance in combination with one or more other types of health insurance. The second insurance-related variable was the percent of individuals without health insurance coverage. Together, these two variables captured populations that were among the least integrated into the U.S. health care system.

**Table e2.2:** Variables calculated from 2015–2019 American Community Survey (ACS) Five-Year Estimates

| Variable | Table | Universe | Numerator | Denominator |
| --- | --- | --- | --- | --- |
| Population | B01001 | Total population | Column 1 | None |
| Population age 15 or older | B01001 | Total population | Sum of columns 6–25 and 30–49 | None |
| Percent age 65 or older | B01001 | Total population | Sum of columns 20–25 and 44–49 | Column 1 |
| Percent employed in health care or social assistance | C24030 | Civilian employed population age 16 or older | Sum of columns 23 and 50 | Column 1 |
| Percent under poverty line | C17002 | Population for whom poverty status is determined | Sum of columns 2–3 | Column 1 |
| Percent with Medicaid or other means-tested public health insurance | B27010 | Civilian non-institutionalized population | Sum of columns 23, 29, 39, 46, and 62 | Column 1 |
| Percent without health insurance | B27010 | Civilian non-institutionalized population | Sum of columns 33, 50, and 66 | Column 1 |
| Percent without internet access | B28002 | Households | Column 13 | Column 1 |
| Percent Black | B03002 | Total population | Column 4 | Column 1 |
| Percent Hispanic | B03002 | Total population | Column 12 | Column 1 |
| Percent Asian | B03002 | Total population | Column 6 | Column 1 |
| Percent White | B03002 | Total population | Column 3 | Column 1 |

We adjusted for income, a key component of SES, by including the percent of the population that was below the poverty line:

> The data on poverty status of households were derived from answers to the income questions. Since poverty is defined at the family level and not the household level, the poverty status of the household is determined by the poverty status of the householder. Households are classified as poor when the total income of the householder’s family is below the appropriate poverty threshold. (For nonfamily householders, their own income is compared with the appropriate threshold.) The income of people living in the household who are unrelated to the householder is not considered when determining the poverty status of a household, nor does their presence affect the family size in determining the appropriate threshold. The poverty thresholds vary depending on three criteria: size of family, number of related children, and, for 1- and 2-person families, age of householder.^14p30^

Percent without internet access was an important independent variable because making appointments online was often the most effective way to secure a vaccine. This variable was the only variable measured at the household level. In the ACS, internet access is defined as “whether or not someone in the household uses or can connect to the internet, regardless of whether or not they pay for the service.”^14p13^ Households are designated as having internet access if at least one member can access the internet through a computer or mobile device.

##### Race/ethnicity

We use the term “race/ethnicity” rather than “race,” “ethnicity,” or “race and ethnicity.” In the 2015–2019 ACS, USCB considered Hispanic, Latino, and Spanish origins as indicators of ethnicity and other origins as indicators of race.^14^ USCB racial/ethnic definitions change over time and are often unaligned with popular understandings of race/ethnicity, academic definitions of race/ethnicity, or analytically appropriate racial/ethnic schemes for a research question or site.^15–22^ A sharp distinction between race and ethnicity does not reflect the processes of stratification in which we were interested in this analysis. The combined term “race/ethnicity” communicates the construct of interest and the structure of the data we used to measure it.

From ACS data,^8, 14^ we created variables measuring the estimated populations of four mutually exclusive, non-exhaustive racial/ethnic groups: Black, Hispanic, Asian, and White. We defined Hispanic as Hispanic, Latino, or Spanish origin, of any race(s). We defined Black, Asian, and White as Black or African American alone, Asian alone, and White alone, respectively, and non-Hispanic. This approach implies a fifth category comprised of non-Hispanic individuals of multiple races or of any other race alone, including American Indians, native Alaskans and Hawaiians, and Pacific Islanders. The four racial/ethnic variables did not sum to one (100 percent) unless the estimated population of the fifth category was zero.

Thoroughly accounting for limitations stemming from USCB racial/ethnic categories was outside the scope of our analysis, but attending to the definitions of racial/ethnic categories aids in interpreting our findings. In the ACS,

> The terms “Hispanic,” “Latino,” and “Spanish” are used interchangeably. Some respondents identify with all three terms while others may identify with only one of these three specific terms. Hispanics or Latinos who identify with the terms “Hispanic,” “Latino,” or “Spanish” are those who classify themselves in one or more of the specific Hispanic, Latino, or Spanish categories listed on the questionnaire (“Mexican,” “Puerto Rican,” or “Cuban”) as well as those who indicate that they are “another Hispanic, Latino, or Spanish origin.” … People who identify their origin as Hispanic, Latino, or Spanish may be of any race.^14p76^

The ACS classifies individuals as White if they report “origins in any of the original peoples of Europe, the Middle East, or North Africa,” including people who “report entries such as Irish, German, Italian, Lebanese, Arab, Moroccan, or Caucasian.”^14p114^ It classifies individuals as Black or African American if they report “origins in any of the Black racial groups of Africa,” including people who “report entries such as African American, Kenyan, Nigerian, or Haitian.”^14p114^ It classifies individuals as Asian if they report “origins in any of the original peoples of the Far East, Southeast Asia, or the Indian subcontinent including, for example, Cambodia, China, India, Japan, Korea, Malaysia, Pakistan, the Philippine Islands, Thailand, and Vietnam.”^14p115^

#### e2.3 Geospatial data

We used several geospatial vector datasets from the USCB 2019 TIGER/Line Shapefiles (TLS). The TLS are extracts of official USCB geographic and cartographic data.^23^ We collected the following TLS data from the USCB API:^24^ city boundaries from the Place State-Based Shapefiles; ZCTA boundaries from the Five-Digit ZCTA National Shapefile; CBG boundaries from the Block Group State-Based Shapefiles; the boundaries of bodies of water from the Area Hydrography County-Based Shapefiles; and the boundaries of USCB-designated landmarks from the Area Landmark State-Based Shapefiles. The coordinate reference system (CRS) for the TLS data was the North American Datum of 1983 (NAD83), an ellipsoidal system that uses geodetic latitude and longitude (not a Cartesian plane).

### e3 Spatial processing

#### e3.1 ZIP Codes, ZIP Code Tabulation Areas (ZCTAs), and vaccination records

For brevity and interpretability, in the main paper we refer to our units of analysis as ZIP Codes, the official and colloquial name for postal codes in the U.S. ZIP Codes were the bases of the units of analysis but themselves were not viable analytical units. ZIP Codes are lists of discrete postal addresses, not areal units. More specifically, they are

> administrative units established by the United States Postal Service (USPS) for the distribution of mail. ZIP Codes serve addresses for the most efficient delivery of mail, and therefore generally do not respect political or census statistical area boundaries. ZIP Codes usually do not have clearly identifiable boundaries, often serve a continually changing area, are changed periodically to meet postal requirements, and do not cover all the land area of the United States.^25pA–13^

**Table e3.1:** Population distribution of ZIP Code Tabulation Areas (ZCTAs) within and across eight large U.S. cities, 2015–2019

| City | ZCTAs | Population |  |  |  | Area |  |  |  | Population density |  |  |  |
| --- | --- | --- | --- | --- | --- | --- | --- | --- | --- | --- | --- | --- | --- |
|  |  | *M* | *SD* | 10% | 90% | *M* | *SD* | 10% | 90% | *M* | *SD* | 10% | 90% |
| New York | 196 | 44,673 | 27,176 | 9,151 | 83,574 | 4.4 | 4.2 | 0.9 | 8.2 | 16,237 | 12,669 | 3,209 | 35,127 |
| Chicago | 87 | 40,687 | 25,148 | 11,559 | 79,690 | 11.8 | 8.5 | 2.6 | 20.2 | 4,825 | 3,617 | 1,378 | 9,804 |
| Houston | 145 | 37,194 | 21,173 | 16,170 | 59,835 | 43.9 | 60.7 | 9.7 | 97.3 | 1,518 | 968 | 410 | 2,789 |
| Phoenix | 67 | 36,025 | 17,375 | 10,064 | 60,657 | 52.1 | 85.6 | 10.4 | 107.5 | 1,500 | 1,004 | 255 | 2,794 |
| Philadelphia | 68 | 28,603 | 18,284 | 6,158 | 53,988 | 9.6 | 7.8 | 3.3 | 17.6 | 4,269 | 3,163 | 863 | 8,371 |
| San Antonio | 69 | 28,439 | 19,865 | 4,818 | 56,665 | 49.1 | 50.1 | 5.8 | 106.7 | 1,043 | 777 | 74 | 2,092 |
| San Diego | 58 | 38,047 | 22,312 | 4,431 | 64,465 | 34.6 | 59.8 | 7.9 | 59.7 | 1,921 | 1,267 | 432 | 3,764 |
| Dallas | 86 | 33,483 | 19,757 | 8,188 | 56,424 | 33.1 | 34.7 | 8.8 | 67.4 | 1,523 | 955 | 467 | 2,629 |
| **Overall** | **776** | **37,495** | **23,203** | **8,754** | **69,876** | **26.6** | **47.8** | **2.1** | **57.5** | **5,835** | **9,016** | **495** | **16,875** |

Includes the total population and geographic area of all ZCTAs that intersected the cities proper. Area measured in square kilometers; corresponds to $A_i$ in Equation e3.2.

We operationalized ZIP Codes as ZCTAs. For each ZIP Code, the USCB delimits a corresponding ZCTA that approximates the ZIP Code as a polygonal areal unit. The area of each ZCTA is mutually exclusive with those of all other ZCTAs.^23^ ZIP Codes and ZCTAs are related and frequently used interchangeably in research, but ZCTAs are suboptimal units for spatial and quantitative analysis.

ZCTAs are distortive and error-prone. ZIP Codes do not have objective, non-overlapping areal boundaries, which often makes converting from ZIP Codes to ZCTAs uncertain. Public health scholars, epidemiologists, and spatial analysts consistently find mismatches between observations’ true locations and recorded ZCTAs.^26–29^ Despite the well-known risk of inducing measurement error, however, most state and local authorities published COVID-19 vaccination data aggregated by ZIP Code. Some failed to indicate whether data were aggregated by ZIP Code or ZCTA. These reporting decisions left researchers without a feasible alternative to using ZCTAs to define areal boundaries consistently across cities.

We summarize the population distribution of each ZCTA that intersected the eight cities proper in Table e3.1. The average ZCTA had 37,495 estimated inhabitants and spanned 26.6 square kilometers, an area nearly half the size of Manhattan or roughly twice the size of Los Angeles International or London Heathrow Airports. In addition, ZCTAs’ physical and population sizes varied widely. Over 20 percent of ZCTAs had fewer than 10,000 or more than 70,000 estimated inhabitants. Some were as small as two square kilometers, just over half the size of Central Park in Manhattan; others approached or exceeded 60 square kilometers, an area larger than that of Manhattan and about one-third the size of Washington, DC.

ZCTAs have much larger geographical areas and populations than colloquial and academic definitions of neighborhoods, which limits their utility for studying inequality. Using ZCTAs as units of analysis complicates making connections with the voluminous literature on neighborhood effects,^30, 31^ which usually uses finer spatial units. For example, census tracts—the most common areal units in analyses of neighborhood inequality in the U.S.—average 4,000 residents and rarely have more than 8,000.^8, 32^ ZIP Codes and ZCTAs also differ substantially from subjective neighborhood boundaries,^33–37^ activity spaces,^38–41^ and other, analytically sounder local units.^42–47^ Aggregating person-level observations to a scale as wide as ZCTAs increases variation within units and decreases variation between them. Statistical analysis may therefore mischaracterize disparities or fail to detect them altogether.^48–53^ For these reasons, we avoid the term “neighborhood” when describing our units and findings, and our study’s evidence of inequality is partial and potentially understated.

Issues of spatial mismatch and scale aside, ZIP Codes and ZCTAs are inconvenient reporting units for comparing outcomes across administrative jurisdictions. ZIP Codes and ZCTAs overlap neighborhoods, municipalities, counties, and states.^8^ As the rightmost column of Table e2.1 shows, the jurisdictions of some agencies that reported vaccination data also overlapped political boundaries, frequently excluding part of the territory of some ZCTAs. Several sources of data extended only to the populations of cities proper, excluding vaccinations administered to residents of other municipalities who lived in ZIP Codes that straddled city borders. Other agencies’ vaccination counts included all individuals with a given ZIP Code, regardless of the municipality in which they resided, within a particular state or county. The administrative and demographic data required adjustment before we could meaningfully compare trends among units.

#### e3.2 Spatial interpolation

To create consistent, comparable units of analysis given reporting irregularities in the vaccination data, we restricted the analysis to the geographical limits of the eight cities proper. We excluded populations residing outside the cities, even if they shared a ZIP Code with some city residents. Because only aggregated administrative and demographic data were available, we relied on spatial interpolation to approximate the within-city values of variables for ZCTAs that spanned city borders.

We used the overlay or areal weighting method of spatial interpolation.^54, 55^ This method estimates the value of a variable in a target zone based on the proportion of smaller source zones that intersect it. Formally,

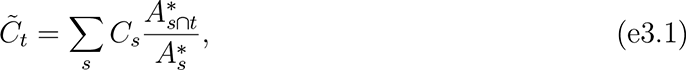

where *C* was the observed value of a count variable; *C̃* was the interpolated value of a count variable; 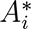 approximated the geographic area of unit *i* that could be populated; 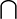 denoted geometric intersection; and *s* and *t* denoted the source and target zones, respectively. We conducted the interpolation using the NAD83 CRS and square kilometers as the areal unit of measure.^56, 57^

The target zones were the geometric intersections between 776 ZCTAs and the eight cities proper. (For ZCTAs that were completely inside city limits, the target zone was the entire ZCTA). The source zones were 16,283 CBGs that intersected the cities proper. The most local areal units for which the USCB releases ACS estimates, CBGs’ populations typically range from 600 to 3,000 people.^8^ The area of 90% of CBGs that intersected the cities was 1.33 square kilometers or less. The typical CBG was 0.93 square kilometers—less than four percent of the area of the typical ZCTA in the study.

The overlay method had several benefits for this study. It also entailed one nontrivial assumption. Overlay interpolation accounts for uneven population density within target zones at the level of source zones. In addition, it is intuitive, computationally inexpensive, and does not require supplemental data. Using the overlay method required us to assume, however, that populations were uniformly distributed within CBGs. This assumption was modest for such small units.

**Table e3.2:**
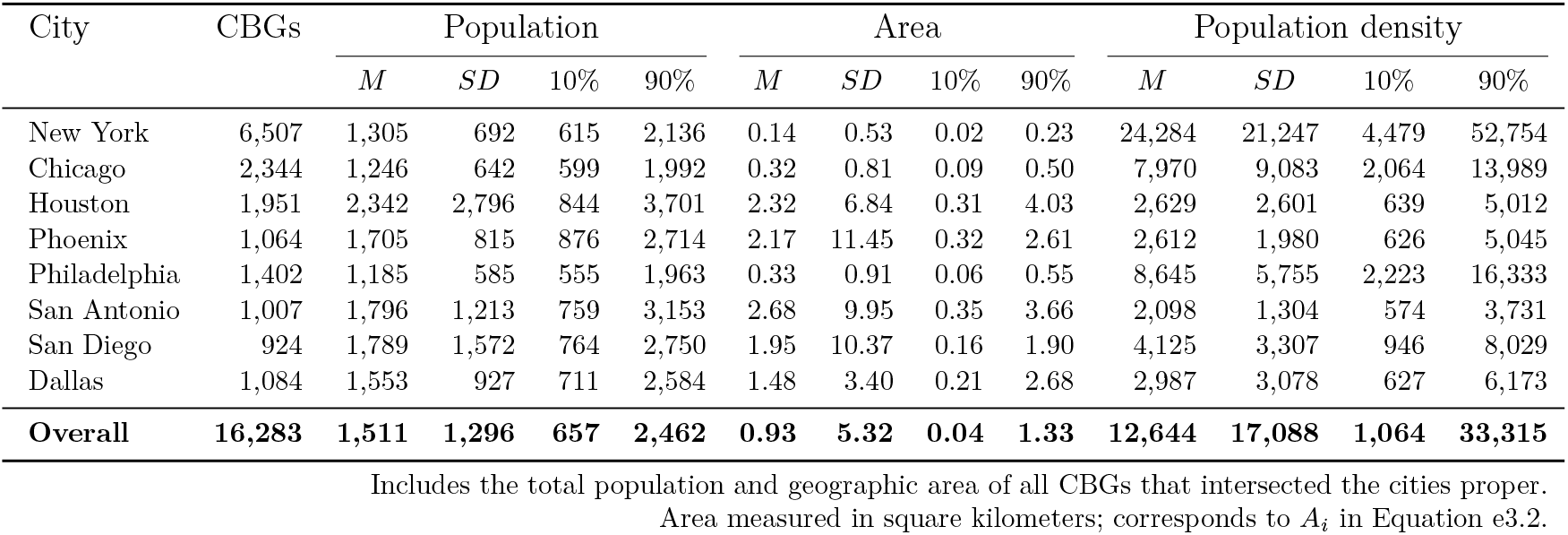
Population distribution of census block groups (CBGs) within and across eight large U.S. cities, 2015–2019

| City | CBGs | Population |  |  |  | Area |  |  |  | Population density |  |  |  |
| --- | --- | --- | --- | --- | --- | --- | --- | --- | --- | --- | --- | --- | --- |
|  |  | *M* | *SD* | 10% | 90% | *M* | *SD* | 10% | 90% | *M* | *SD* | 10% | 90% |
| New York | 6,507 | 1,305 | 692 | 615 | 2,136 | 0.14 | 0.53 | 0.02 | 0.23 | 24,284 | 21,247 | 4,479 | 52,754 |
| Chicago | 2,344 | 1,246 | 642 | 599 | 1,992 | 0.32 | 0.81 | 0.09 | 0.50 | 7,970 | 9,083 | 2,064 | 13,989 |
| Houston | 1,951 | 2,342 | 2,796 | 844 | 3,701 | 2.32 | 6.84 | 0.31 | 4.03 | 2,629 | 2,601 | 639 | 5,012 |
| Phoenix | 1,064 | 1,705 | 815 | 876 | 2,714 | 2.17 | 11.45 | 0.32 | 2.61 | 2,612 | 1,980 | 626 | 5,045 |
| Philadelphia | 1,402 | 1,185 | 585 | 555 | 1,963 | 0.33 | 0.91 | 0.06 | 0.55 | 8,645 | 5,755 | 2,223 | 16,333 |
| San Antonio | 1,007 | 1,796 | 1,213 | 759 | 3,153 | 2.68 | 9.95 | 0.35 | 3.66 | 2,098 | 1,304 | 574 | 3,731 |
| San Diego | 924 | 1,789 | 1,572 | 764 | 2,750 | 1.95 | 10.37 | 0.16 | 1.90 | 4,125 | 3,307 | 946 | 8,029 |
| Dallas | 1,084 | 1,553 | 927 | 711 | 2,584 | 1.48 | 3.40 | 0.21 | 2.68 | 2,987 | 3,078 | 627 | 6,173 |
| **Overall** | **16,283** | **1,511** | **1,296** | **657** | **2,462** | **0.93** | **5.32** | **0.04** | **1.33** | **12,644** | **17,088** | **1,064** | **33,315** |

Includes the total population and geographic area of all CBGs that intersected the cities proper. Area measured in square kilometers; corresponds to $A_i$ in Equation e3.2.

We nonetheless took steps to mitigate potential inaccuracies. As Equation e3.1 indicates, we interpolated 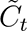 from *A^∗^* rather than from *A*. Formally,

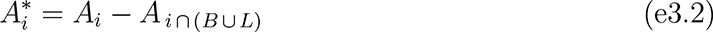

where *A_i_* was the total area of unit *i*; ∪ denoted geometric union; *B* was the set of USCB-designated bodies of water; and *L* was a subset of USCB-designated landmarks. *B* and *L* overlapped in many locations. *B* included the area of “perennial and intermittent … ponds, lakes, oceans, swamps, glaciers, and … large streams.”^23p3-34^ We list the types of landmarks included in *L* in Table e3.3. By excluding from areal calculations parts of source zones that were unlikely to contribute population to the target zones, the adjustment in Equation e3.2 made it more plausible to assume uniform population density within CBGs.

We visualize the overlay interpolation process in Figure e3.1. We used overlay interpolation to estimate the target zones’ numerators and denominators for all variables listed in Table e2.2—including the population age 15 and older, the outcome variable’s denominator.

Calculating the numerator for the outcome variable—the number of individuals with at least dose of a COVID-19 vaccine—required a separate interpolation procedure. For New York, Chicago, and Philadelphia, agencies’ counts included only vaccinated individuals living inside the cities proper. In these cases, we could adopt the reported total directly as the numerator for the outcome variable. Elsewhere, reporting agencies’ counts for ZIP Codes intersecting the cities included vaccinated individuals residing outside the cities proper. We resolved this discrepancy by adjusting the reported counts using multipler *m*. The value of *m* was one for observations in New York, Chicago, and Philadelphia; otherwise, it was the ratio between the estimated populations of the target zone and its corresponding reporting unit. Formally,

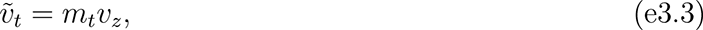

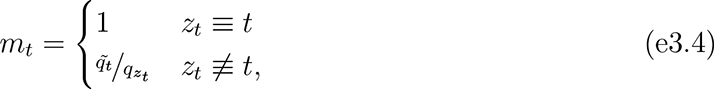

where *v_i_* and *ṽ_i_* were the observed and interpolated numbers of individuals with at least one vaccine dose in unit *i*, respectively; *q_i_* and *q̃_i_* were the observed and interpolated total populations of unit *i*, respectively; and *z_t_* denoted the area covered by the reporting unit corresponding to target zone *t*. This adjustment assumed that, within ZCTAs that straddled city borders, vaccinated individuals were distributed proportionally to the total population.

**Table e3.3:** U.S. Census Bureau (USCB)-designated landmarks excluded from area calculations

| Code | Description |
| --- | --- |
| C3026 | Quarry or mine |
| C3077 | Solar farm |
| K2180 | Park |
| K2181 | National park, forest, etc. |
| K2182 | Other federal land |
| K2183 | Tribal park, forest, etc. |
| K2184 | State park, forest, etc. |
| K2185 | Regional park, forest, etc. |
| K2186 | County park, forest, etc. |
| K2187 | Civil division park, forest, etc. |
| K2188 | Municipal park, forest, etc. |
| K2189 | Private park, forest, etc. |
| K2190 | Other park, forest, etc. |
| K2362 | Industrial building or park |
| K2424 | Marina |
| K2432 | Pier or dock |
| K2451 | Airport |
| K2452 | Train station |
| K2453 | Bus terminal |
| K2454 | Marine terminal |
| K2455 | Seaplane anchorage |
| K2456 | Airport terminals |
| K2457 | Airport grounds |
| K2458 | Park-and-ride facility |
| K2459 | Airport runway |
| K2460 | Helicopter landing |
| K2561 | Golf course |
| K2564 | Amusement park |
| K2582 | Cemetery |
| K2586 | Zoo |

Codes are USCB MAF/TIGER Feature Class Codes<sup>23</sup>.

**Figure e3.1:**
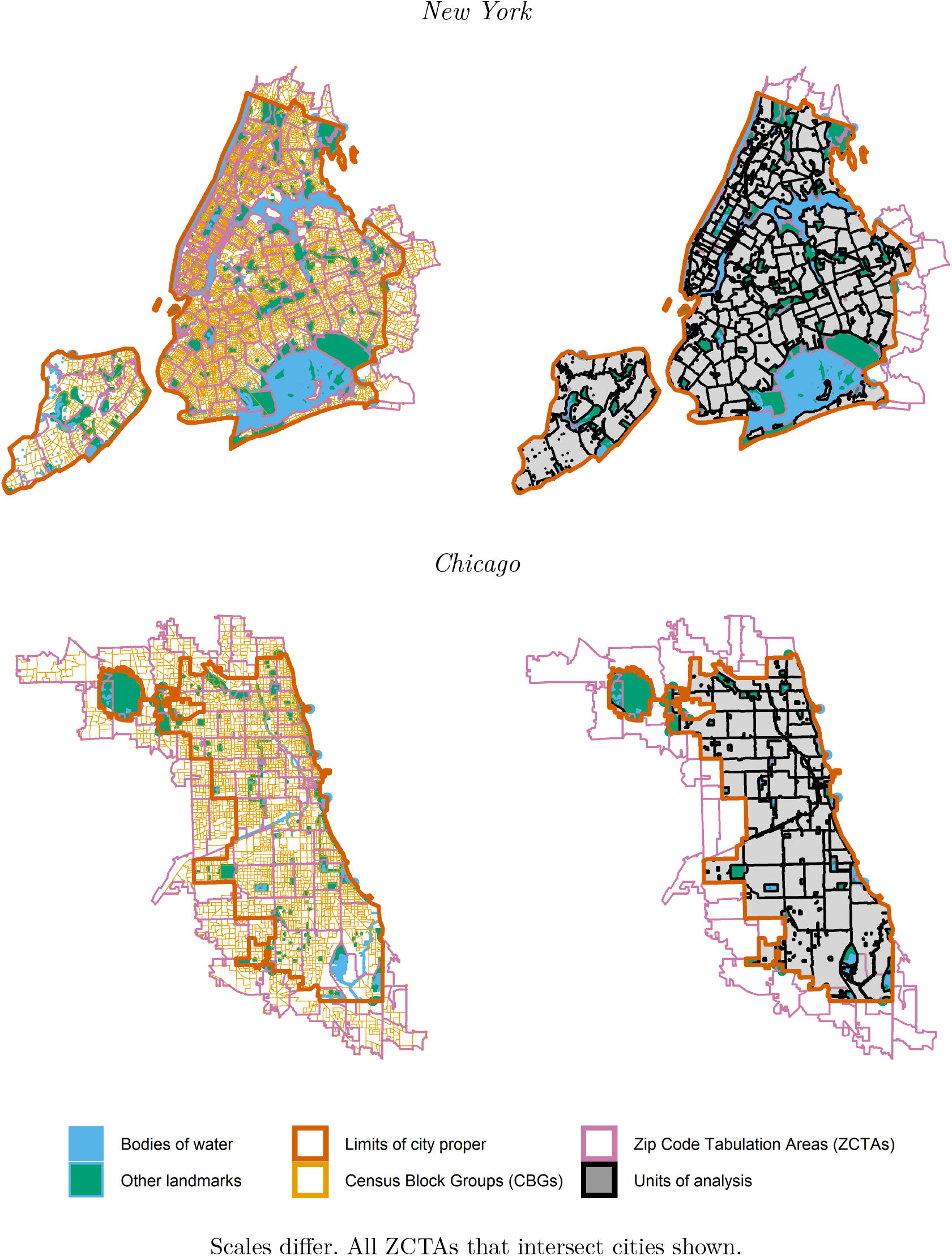
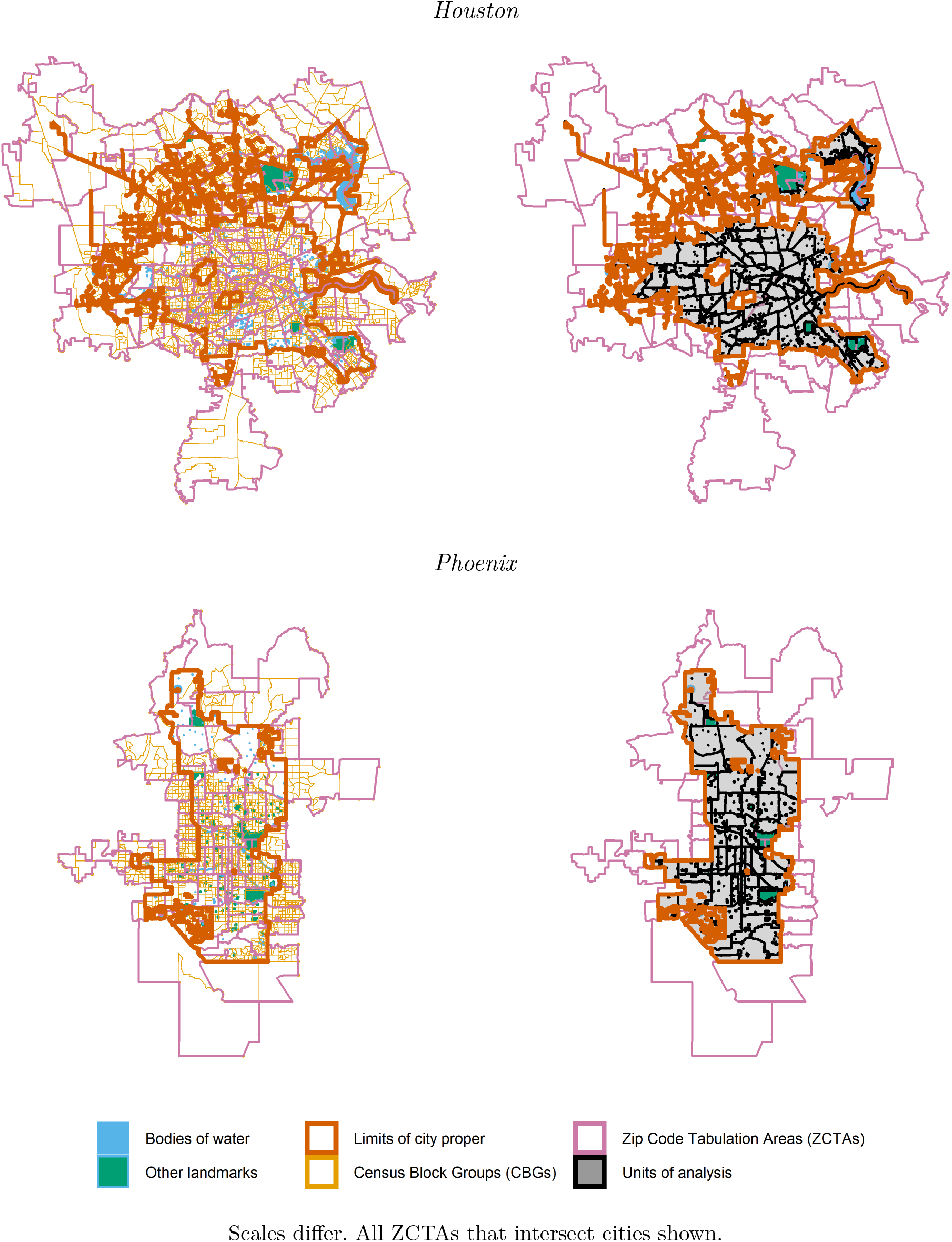
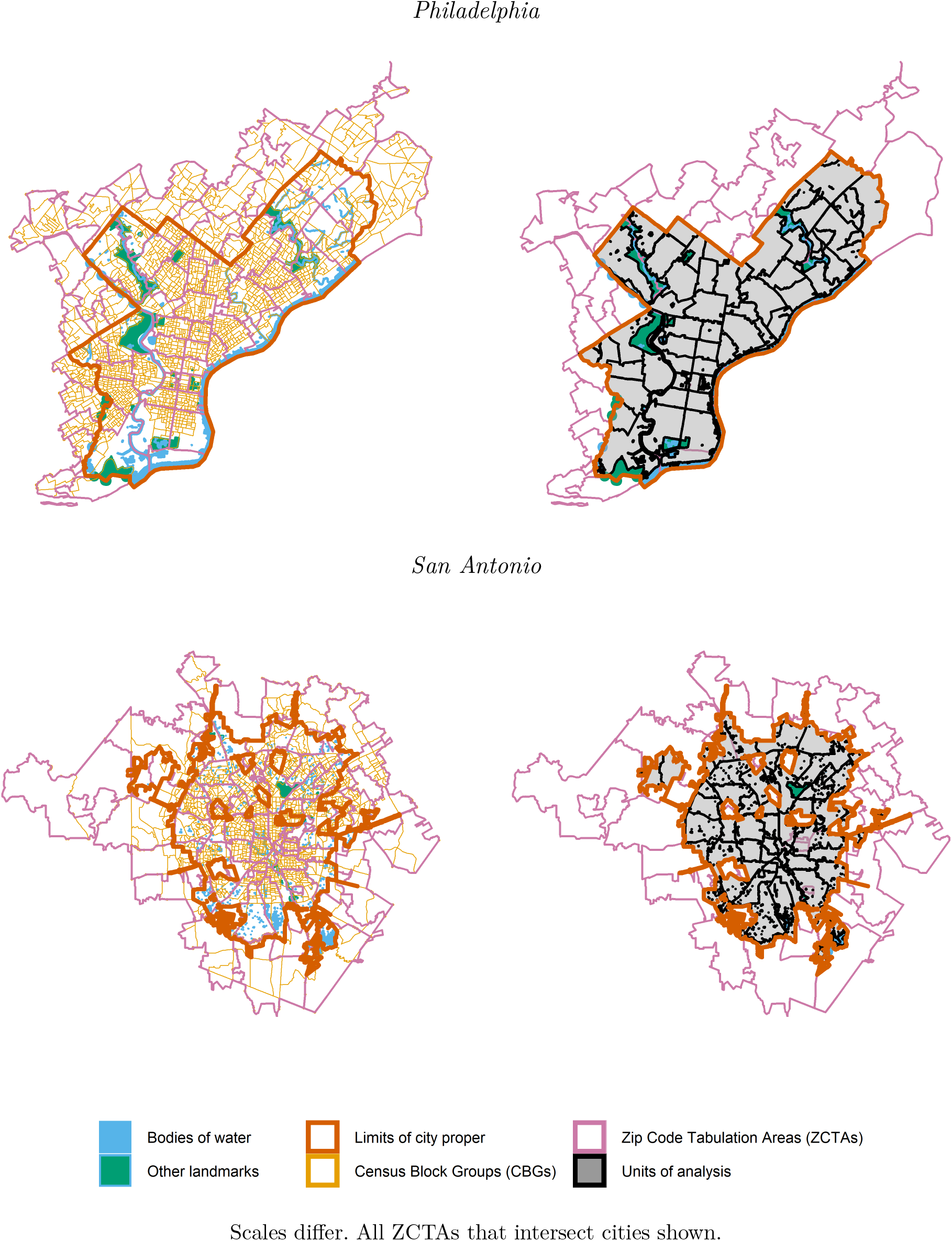
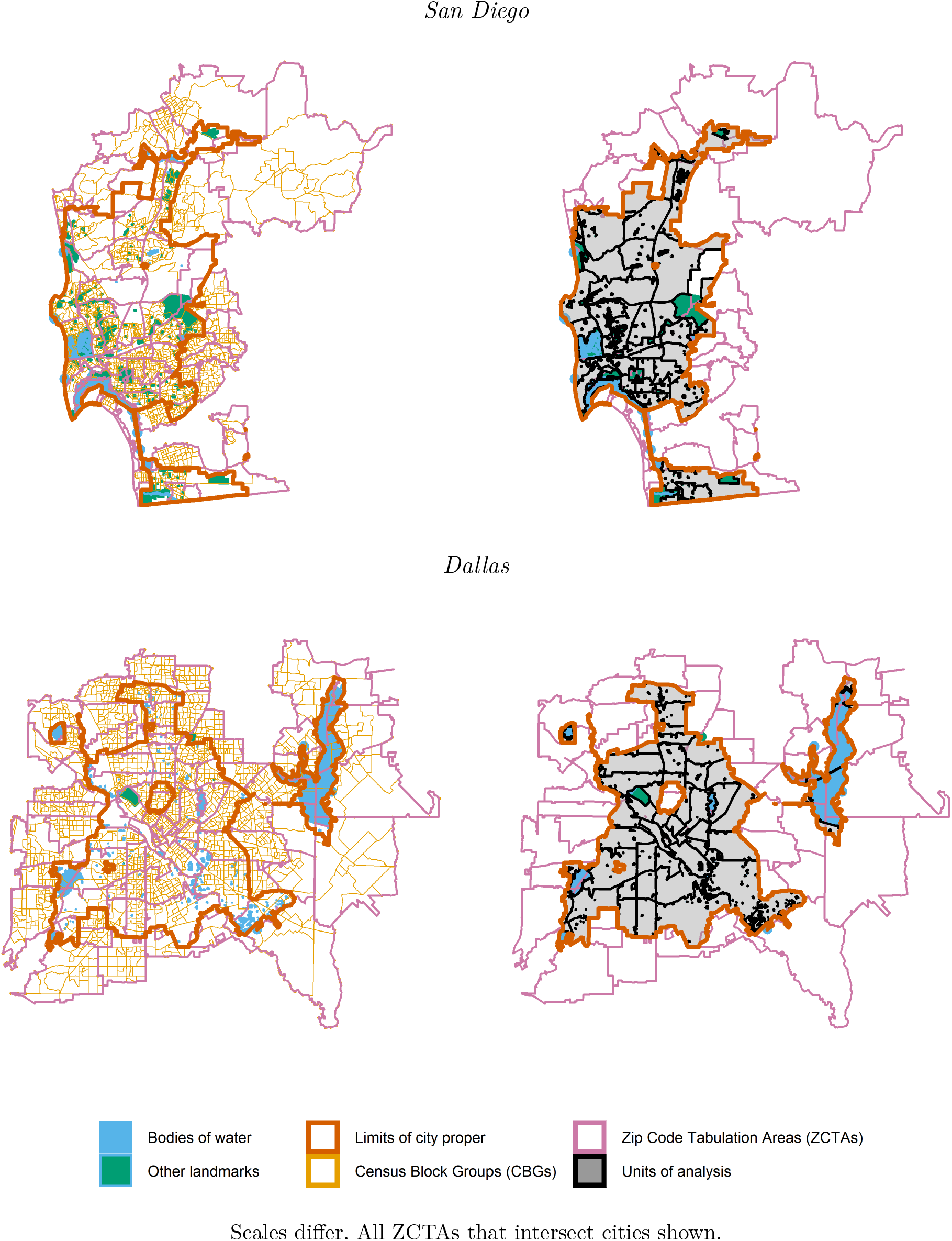
Summary of overlay interpolation by city

**Table e3.4:** Population distribution of interpolated units of analysis within and across eight large U.S. cities, 2015–2019

| City | Units | Population |  |  |  | Area |  |  |  | Population density |  |  |  |
| --- | --- | --- | --- | --- | --- | --- | --- | --- | --- | --- | --- | --- | --- |
|  |  | *M* | *SD* | 10% | 90% | *M* | *SD* | 10% | 90% | *M* | *SD* | 10% | 90% |
| New York | 175 | 48,078 | 26,244 | 18,050 | 84,458 | 3.6 | 3.3 | 1.0 | 6.2 | 18,864 | 12,726 | 5,443 | 36,182 |
| Chicago | 53 | 51,131 | 24,084 | 21,882 | 86,091 | 9.7 | 8.3 | 2.2 | 16.6 | 7,376 | 4,215 | 3,271 | 13,798 |
| Houston | 99 | 24,998 | 15,270 | 6,933 | 43,964 | 16.0 | 9.1 | 5.4 | 28.4 | 1,708 | 928 | 745 | 2,830 |
| Phoenix | 50 | 32,331 | 17,292 | 7,825 | 53,528 | 25.7 | 20.6 | 5.6 | 54.0 | 1,707 | 990 | 360 | 3,035 |
| Philadelphia | 46 | 34,328 | 17,463 | 12,436 | 59,943 | 7.1 | 4.3 | 2.4 | 13.6 | 5,965 | 2,899 | 2,295 | 9,673 |
| San Antonio | 48 | 31,560 | 17,439 | 6,984 | 56,254 | 25.9 | 15.4 | 7.1 | 48.1 | 1,392 | 704 | 482 | 2,360 |
| San Diego | 33 | 42,400 | 18,163 | 22,666 | 67,742 | 22.1 | 16.4 | 8.8 | 46.8 | 2,603 | 1,487 | 947 | 4,690 |
| Dallas | 48 | 27,830 | 20,596 | 4,760 | 56,793 | 18.2 | 15.5 | 4.1 | 38.7 | 1,857 | 1,035 | 734 | 3,592 |
| **Overall** | **552** | **38,123** | **23,314** | **9,166** | **72,696** | **13.0** | **13.8** | **1.7** | **31.1** | **8,085** | **10,555** | **901** | **22,545** |

Area measured in square kilometers; corresponds to $A_t^*$ in Equation e3.2.

We made two final adjustments to the interpolated units of analysis. The interpolation yielded several target zones with very small estimated populations. To minimize the effects of outliers and reduce variation in the size of the units, we merged target zones with unusually low populations into adjacent zones such that no unit of analysis had fewer than 3,000 estimated residents. In addition, in a small number cases, implausible or extreme values surfaced in the interpolated vaccination outcome variables. These values suggested spatial mismatch occurred in the ZIP Code-level data, or that the interpolation was particularly ill-suited to the spatial contours of a portion of the study area. To smooth out mismatches and discrepancies in these instances, we merged sets of contiguous zones that visual inspection suggested were susceptible to administrative mismatch or inaccurate interpolation. Due to these adjustments, there were 776 target zones but only 552 final units of analysis.

We summarize the population distribution of the 552 ZIP Code-based units of analysis in Table e3.4, and we illustrate them in the right-hand panels of Figure e3.1. The average unit of analysis had an interpolated total population of 38,123, roughly equivalent to the average population of the ZCTAs that were the bases of the target zones. Units’ populations varied considerably. About one-quarter had more than 50,000 estimated inhabitants; another quarter had fewer than 20,000. Because the units excluded bodies of water, landmarks, and area outside city limits, however, the average physical size of the units of analysis was roughly half that of the average ZCTA. Furthermore, the area of the units of analysis exhibited significantly less variation than the raw ZCTAs. On the whole, the interpolated spatial units provided more viable bases for analysis and comparison than were available in the raw data. Yet they generally remained susceptible to the analytical pitfalls of ZIP Codes and ZCTAs that we discuss in Section e3.1. We present descriptive statistics for observations on the analysis variables for the interpolated units of analysis in Table e3.6.

**Table e3.5:** Official vs. interpolated estimates of the populations of eight large U.S. cities, 2015–2019

|  | Official | Interpolated | Difference |
| --- | --- | --- | --- |
| New York | 8,419,316 | 8,413,650 | −0.1% |
| Chicago | 2,709,534 | 2,709,943 | +0.0% |
| Houston | 2,310,432 | 2,474,802 | +7.1% |
| Phoenix | 1,633,017 | 1,616,550 | −1.0% |
| Philadelphia | 1,579,075 | 1,579,088 | +0.0% |
| San Antonio | 1,508,083 | 1,514,880 | +0.5% |
| San Diego | 1,409,573 | 1,399,200 | −0.7% |
| Dallas | 1,330,612 | 1,335,840 | +0.4% |
| **Overall** | **20,899,642** | **21,043,953** | **+0.7%** |

Official estimates from ACS (2015–2019) table B01001, column 1.

#### e3.3 Measurement error and information bias

In regression analysis, information bias arises when variables are measured inaccurately.^58^ If outcome variables are measured with error, regression coefficients are unbiased but confidence intervals too wide.^59, 60^ Researchers might consequently accept null hypotheses they should reject. If independent variables are measured with error, the direction of information bias is case-specific, determined by the covariance of the variables in question.^59, 61^ Typically, regression coefficients are biased towards zero when independent variables are measured with error.^60, 62^ As a result, researchers are vulnerable to understating the magnitude of associations. Measurement error often makes regression inference more conservative.

The nature of available data for this study made some information bias likely. The demographic data were estimates with margins of error.^8^ The administrative records were vulnerable to spatial mismatch between individuals’ ZIP Codes and assigned ZCTAs. Even careful variable construction from these data also probably introduced some measurement error because available quantities were not always aligned with the theoretical quantity of interest. Similarly, as an estimation process based on aggregated observations, overlay interpolation contributed some measurement error.

It is unlikely that these sources of information bias seriously jeopardized our findings. The direction and magnitude of bias from measurement error are difficult to predict, but information bias probably only made our analysis more cautious. The quality of the data was high overall. We used established methods to calculate variables that tracked closely with theoretical variables of interest. Moreover, because information bias typically makes regression findings more conservative, false negatives (Type II errors) were more likely than false positives (Type I errors). The most plausible effect of information bias in this study was to compound the effects of spatial scale that we discuss in Section e3.1: underestimating associations and disparities.

**Table e3.6:**
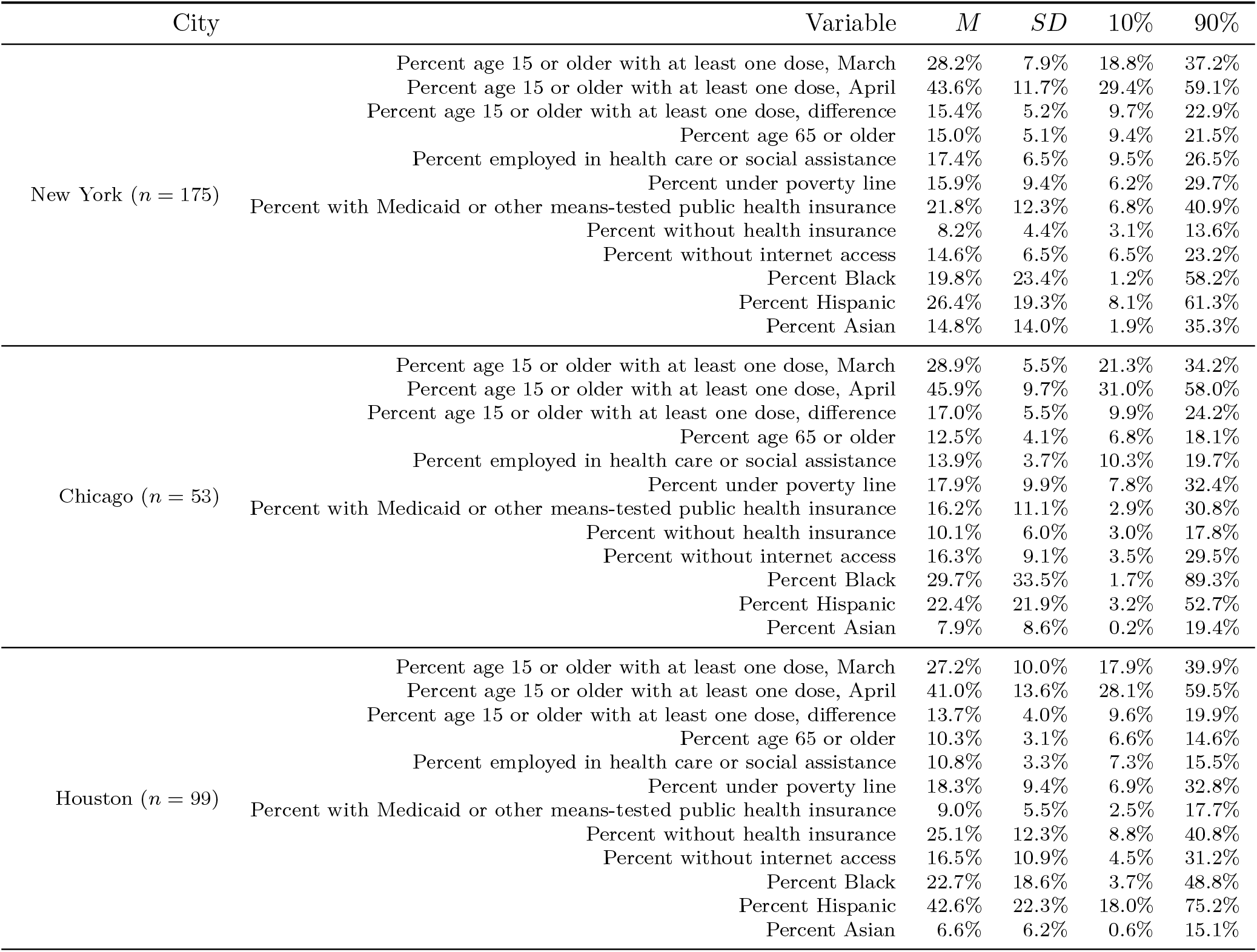
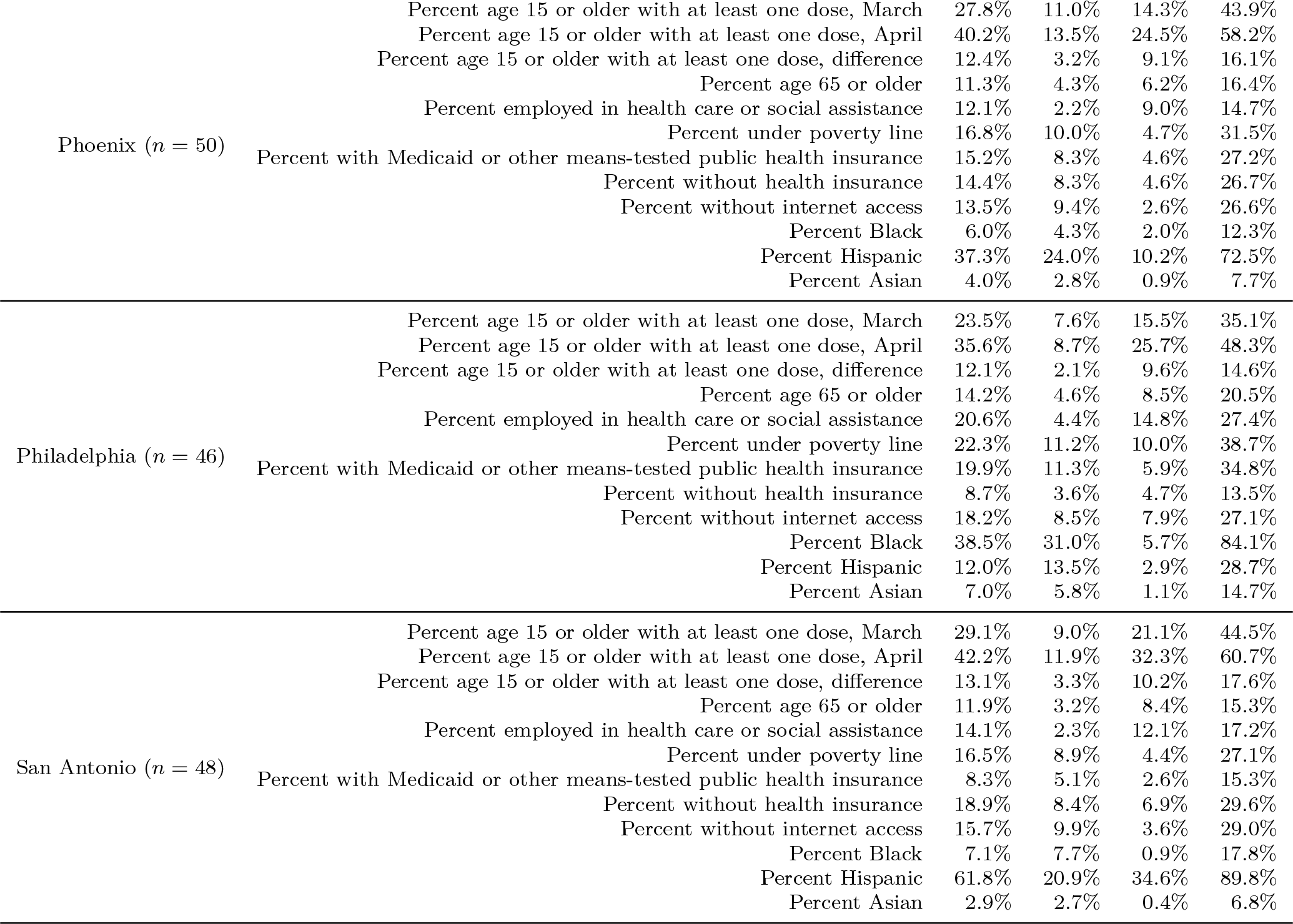
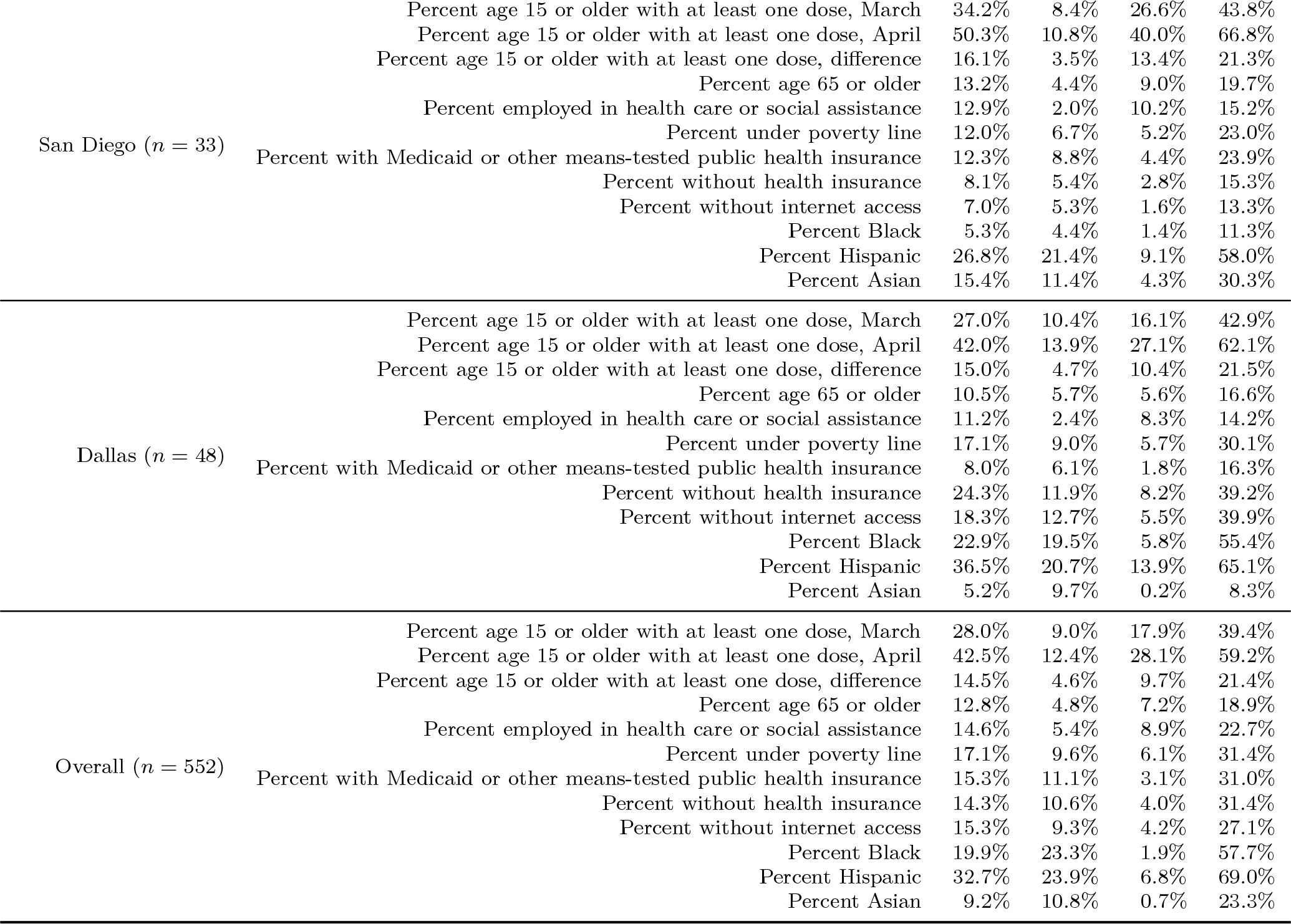
Descriptive statistics on COVID-19 vaccination and population composition in interpolated units of analysis within and across eight large U.S. cities, March and April 2021 (expanded)

| City | Variable | M | SD | 10% | 90% |
| --- | --- | --- | --- | --- | --- |
| New York (*n* = 175) | Percent age 15 or older with at least one dose, March | 28.2% | 7.9% | 18.8% | 37.2% |
|  | Percent age 15 or older with at least one dose, April | 43.6% | 11.7% | 29.4% | 59.1% |
|  | Percent age 15 or older with at least one dose, difference | 15.4% | 5.2% | 9.7% | 22.9% |
|  | Percent age 65 or older | 15.0% | 5.1% | 9.4% | 21.5% |
|  | Percent employed in health care or social assistance | 17.4% | 6.5% | 9.5% | 26.5% |
|  | Percent under poverty line | 15.9% | 9.4% | 6.2% | 29.7% |
|  | Percent with Medicaid or other means-tested public health insurance | 21.8% | 12.3% | 6.8% | 40.9% |
|  | Percent without health insurance | 8.2% | 4.4% | 3.1% | 13.6% |
|  | Percent without internet access | 14.6% | 6.5% | 6.5% | 23.2% |
|  | Percent Black | 19.8% | 23.4% | 1.2% | 58.2% |
|  | Percent Hispanic | 26.4% | 19.3% | 8.1% | 61.3% |
|  | Percent Asian | 14.8% | 14.0% | 1.9% | 35.3% |
| Chicago (*n* = 53) | Percent age 15 or older with at least one dose, March | 28.9% | 5.5% | 21.3% | 34.2% |
|  | Percent age 15 or older with at least one dose, April | 45.9% | 9.7% | 31.0% | 58.0% |
|  | Percent age 15 or older with at least one dose, difference | 17.0% | 5.5% | 9.9% | 24.2% |
|  | Percent age 65 or older | 12.5% | 4.1% | 6.8% | 18.1% |
|  | Percent employed in health care or social assistance | 13.9% | 3.7% | 10.3% | 19.7% |
|  | Percent under poverty line | 17.9% | 9.9% | 7.8% | 32.4% |
|  | Percent with Medicaid or other means-tested public health insurance | 16.2% | 11.1% | 2.9% | 30.8% |
|  | Percent without health insurance | 10.1% | 6.0% | 3.0% | 17.8% |
|  | Percent without internet access | 16.3% | 9.1% | 3.5% | 29.5% |
|  | Percent Black | 29.7% | 33.5% | 1.7% | 89.3% |
|  | Percent Hispanic | 22.4% | 21.9% | 3.2% | 52.7% |
|  | Percent Asian | 7.9% | 8.6% | 0.2% | 19.4% |
| Houston (*n* = 99) | Percent age 15 or older with at least one dose, March | 27.2% | 10.0% | 17.9% | 39.9% |
|  | Percent age 15 or older with at least one dose, April | 41.0% | 13.6% | 28.1% | 59.5% |
|  | Percent age 15 or older with at least one dose, difference | 13.7% | 4.0% | 9.6% | 19.9% |
|  | Percent age 65 or older | 10.3% | 3.1% | 6.6% | 14.6% |
|  | Percent employed in health care or social assistance | 10.8% | 3.3% | 7.3% | 15.5% |
|  | Percent under poverty line | 18.3% | 9.4% | 6.9% | 32.8% |
|  | Percent with Medicaid or other means-tested public health insurance | 9.0% | 5.5% | 2.5% | 17.7% |
|  | Percent without health insurance | 25.1% | 12.3% | 8.8% | 40.8% |
|  | Percent without internet access | 16.5% | 10.9% | 4.5% | 31.2% |
|  | Percent Black | 22.7% | 18.6% | 3.7% | 48.8% |
|  | Percent Hispanic | 42.6% | 22.3% | 18.0% | 75.2% |
|  | Percent Asian | 6.6% | 6.2% | 0.6% | 15.1% |
| Phoenix ($n = 50$) | Percent age 15 or older with at least one dose, March | 27.8% | 11.0% | 14.3% | 43.9% |
|  | Percent age 15 or older with at least one dose, April | 40.2% | 13.5% | 24.5% | 58.2% |
|  | Percent age 15 or older with at least one dose, difference | 12.4% | 3.2% | 9.1% | 16.1% |
|  | Percent age 65 or older | 11.3% | 4.3% | 6.2% | 16.4% |
|  | Percent employed in health care or social assistance | 12.1% | 2.2% | 9.0% | 14.7% |
|  | Percent under poverty line | 16.8% | 10.0% | 4.7% | 31.5% |
|  | Percent with Medicaid or other means-tested public health insurance | 15.2% | 8.3% | 4.6% | 27.2% |
|  | Percent without health insurance | 14.4% | 8.3% | 4.6% | 26.7% |
|  | Percent without internet access | 13.5% | 9.4% | 2.6% | 26.6% |
|  | Percent Black | 6.0% | 4.3% | 2.0% | 12.3% |
|  | Percent Hispanic | 37.3% | 24.0% | 10.2% | 72.5% |
|  | Percent Asian | 4.0% | 2.8% | 0.9% | 7.7% |
| Philadelphia ($n = 46$) | Percent age 15 or older with at least one dose, March | 23.5% | 7.6% | 15.5% | 35.1% |
|  | Percent age 15 or older with at least one dose, April | 35.6% | 8.7% | 25.7% | 48.3% |
|  | Percent age 15 or older with at least one dose, difference | 12.1% | 2.1% | 9.6% | 14.6% |
|  | Percent age 65 or older | 14.2% | 4.6% | 8.5% | 20.5% |
|  | Percent employed in health care or social assistance | 20.6% | 4.4% | 14.8% | 27.4% |
|  | Percent under poverty line | 22.3% | 11.2% | 10.0% | 38.7% |
|  | Percent with Medicaid or other means-tested public health insurance | 19.9% | 11.3% | 5.9% | 34.8% |
|  | Percent without health insurance | 8.7% | 3.6% | 4.7% | 13.5% |
|  | Percent without internet access | 18.2% | 8.5% | 7.9% | 27.1% |
|  | Percent Black | 38.5% | 31.0% | 5.7% | 84.1% |
|  | Percent Hispanic | 12.0% | 13.5% | 2.9% | 28.7% |
|  | Percent Asian | 7.0% | 5.8% | 1.1% | 14.7% |
| San Antonio ($n = 48$) | Percent age 15 or older with at least one dose, March | 29.1% | 9.0% | 21.1% | 44.5% |
|  | Percent age 15 or older with at least one dose, April | 42.2% | 11.9% | 32.3% | 60.7% |
|  | Percent age 15 or older with at least one dose, difference | 13.1% | 3.3% | 10.2% | 17.6% |
|  | Percent age 65 or older | 11.9% | 3.2% | 8.4% | 15.3% |
|  | Percent employed in health care or social assistance | 14.1% | 2.3% | 12.1% | 17.2% |
|  | Percent under poverty line | 16.5% | 8.9% | 4.4% | 27.1% |
|  | Percent with Medicaid or other means-tested public health insurance | 8.3% | 5.1% | 2.6% | 15.3% |
|  | Percent without health insurance | 18.9% | 8.4% | 6.9% | 29.6% |
|  | Percent without internet access | 15.7% | 9.9% | 3.6% | 29.0% |
|  | Percent Black | 7.1% | 7.7% | 0.9% | 17.8% |
|  | Percent Hispanic | 61.8% | 20.9% | 34.6% | 89.8% |
|  | Percent Asian | 2.9% | 2.7% | 0.4% | 6.8% |

| City | Variable | *M* | *SD* | 10% | 90% |
| --- | --- | --- | --- | --- | --- |
| San Diego (*n* = 33) | Percent age 15 or older with at least one dose, March | 34.2% | 8.4% | 26.6% | 43.8% |
|  | Percent age 15 or older with at least one dose, April | 50.3% | 10.8% | 40.0% | 66.8% |
|  | Percent age 15 or older with at least one dose, difference | 16.1% | 3.5% | 13.4% | 21.3% |
|  | Percent age 65 or older | 13.2% | 4.4% | 9.0% | 19.7% |
|  | Percent employed in health care or social assistance | 12.9% | 2.0% | 10.2% | 15.2% |
|  | Percent under poverty line | 12.0% | 6.7% | 5.2% | 23.0% |
|  | Percent with Medicaid or other means-tested public health insurance | 12.3% | 8.8% | 4.4% | 23.9% |
|  | Percent without health insurance | 8.1% | 5.4% | 2.8% | 15.3% |
|  | Percent without internet access | 7.0% | 5.3% | 1.6% | 13.3% |
|  | Percent Black | 5.3% | 4.4% | 1.4% | 11.3% |
|  | Percent Hispanic | 26.8% | 21.4% | 9.1% | 58.0% |
|  | Percent Asian | 15.4% | 11.4% | 4.3% | 30.3% |
| Dallas (*n* = 48) | Percent age 15 or older with at least one dose, March | 27.0% | 10.4% | 16.1% | 42.9% |
|  | Percent age 15 or older with at least one dose, April | 42.0% | 13.9% | 27.1% | 62.1% |
|  | Percent age 15 or older with at least one dose, difference | 15.0% | 4.7% | 10.4% | 21.5% |
|  | Percent age 65 or older | 10.5% | 5.7% | 5.6% | 16.6% |
|  | Percent employed in health care or social assistance | 11.2% | 2.4% | 8.3% | 14.2% |
|  | Percent under poverty line | 17.1% | 9.0% | 5.7% | 30.1% |
|  | Percent with Medicaid or other means-tested public health insurance | 8.0% | 6.1% | 1.8% | 16.3% |
|  | Percent without health insurance | 24.3% | 11.9% | 8.2% | 39.2% |
|  | Percent without internet access | 18.3% | 12.7% | 5.5% | 39.9% |
|  | Percent Black | 22.9% | 19.5% | 5.8% | 55.4% |
|  | Percent Hispanic | 36.5% | 20.7% | 13.9% | 65.1% |
|  | Percent Asian | 5.2% | 9.7% | 0.2% | 8.3% |
| Overall (*n* = 552) | Percent age 15 or older with at least one dose, March | 28.0% | 9.0% | 17.9% | 39.4% |
|  | Percent age 15 or older with at least one dose, April | 42.5% | 12.4% | 28.1% | 59.2% |
|  | Percent age 15 or older with at least one dose, difference | 14.5% | 4.6% | 9.7% | 21.4% |
|  | Percent age 65 or older | 12.8% | 4.8% | 7.2% | 18.9% |
|  | Percent employed in health care or social assistance | 14.6% | 5.4% | 8.9% | 22.7% |
|  | Percent under poverty line | 17.1% | 9.6% | 6.1% | 31.4% |
|  | Percent with Medicaid or other means-tested public health insurance | 15.3% | 11.1% | 3.1% | 31.0% |
|  | Percent without health insurance | 14.3% | 10.6% | 4.0% | 31.4% |
|  | Percent without internet access | 15.3% | 9.3% | 4.2% | 27.1% |
|  | Percent Black | 19.9% | 23.3% | 1.9% | 57.7% |
|  | Percent Hispanic | 32.7% | 23.9% | 6.8% | 69.0% |
|  | Percent Asian | 9.2% | 10.8% | 0.7% | 23.3% |

Table e3.5 provides a partial assessment of the level of measurement error stemming from spatial interpolation. The table compares the official ACS population estimates with the sums of the interpolated populations of the target zones. For all cities except Houston and for the eight cities combined, the interpolated population was within one percent of the official estimate. The much greater difference between the estimates for Houston is probably attributable to its irregular physical boundaries, as illustrated in Figure e3.1. This layout would be challenging for any method of spatial interpolation. On average, however, Table e3.5 suggests our overlay interpolation procedure distributed populations with considerable accuracy. This comparison is insufficient to rule out information bias from the interpolation, but it suggests the biasing effects of resulting measurement error were minor overall.

### e4 Analytical approach

#### e4.1 Model specification and estimation

We estimated standard linear models (SLMs) by weighted least squares (LS) and spatial error models (SEMs) with nearest-neighbor spatial weights by maximum likelihood.^63–67^ Formally, for observations of *n* = 552 units on *ρ* independent variables and *k* = 8 nearest neighbors, the SLM was

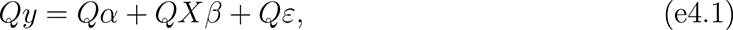

and the SEM was

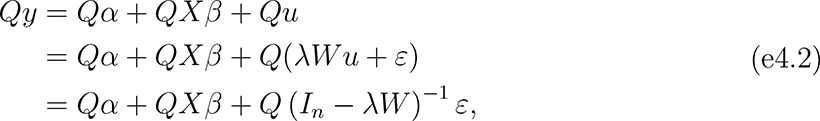

with

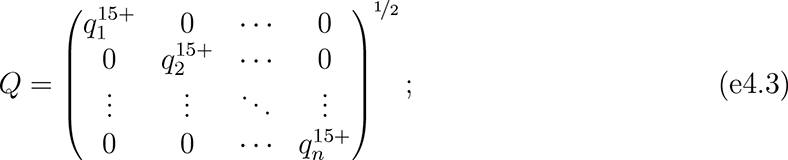

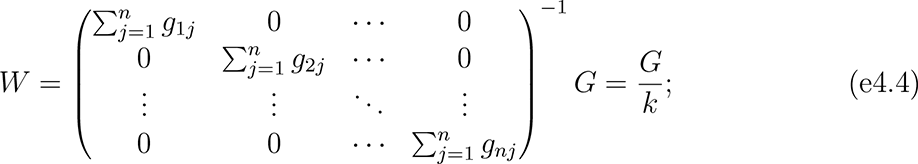

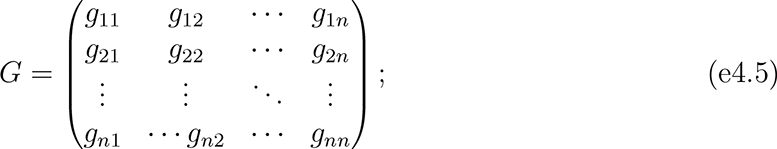

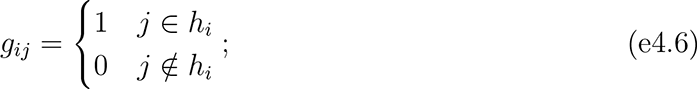

where *y* was an *n* × 1 vector of observations on the outcome variable; *X* was an *n × ρ* matrix of observations on the independent variables; *I_n_* was an identity matrix of size *n*; 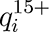 was the estimated population age 15 and older of unit *i*; *h_i_* indexed the *k* units closest to unit *i*; *α* was the intercept (constant) parameter to be estimated; *β* was a 1 × *ρ* vector of coefficients to be estimated; *ε* was an *n* × 1 vector of disturbances; *λ* was a scalar parameter to be estimated measuring average spatial autocorrelation in *ε*, conditional on *W* ; and population weights matrix *Q*, spatial weights matrix *W*, and spatial links matrix *G* were square, of order *n*.

We report expanded step-wise results of both sets of models in Tables e4.1 and e4.2, and we compare the estimates between the SEMs and SLMs that included all independent variables in Figure e4.1. For the March and April outcomes, the coefficient estimates differed significantly or very nearly significantly between the models for three of the SES indicators. The magnitudes of the coefficients for the insurance-related variables decreased from the SLM to the SEM but remained negative, while the coefficient for the poverty variable changed from slightly positive to slightly negative.

#### e4.2 Spatial heterogeneity and modeling strategy

##### Testing for and modeling spatial effects

Estimating SLMs by LS, the most common tool for analyzing high-dimensional relationships among variables, is unbiased and efficient in many settings.^64^ In the presence of spatial effects, however, LS estimates are inaccurate and/or require larger samples to model relationships.^65, 66, 68, 69^ There are two basic classes of spatial effects: spatial heterogeneity and spatial dependence.

Spatial heterogeneity “is related to the lack of stability over space of the … relationships under study. More precisely, this implies that functional forms and parameters vary with location and are not homogeneous throughout the data set.”^65p9^ It was plausible that the relationships among vaccination, priority population composition, socioeconomic composition, and racial/ethnic composition varied from unit to unit but were spatially clustered, even within cities. In principle, the likely sources of spatial heterogeneity could be modeled as independent variables; in practice, they were unobserved. Potential unmeasurable sources of spatial heterogeneity included past levels of exposure to COVID-19, hyperlocal idiosyncrasies in the effects or implementation of vaccination policies, and cultural influences.

Spatial dependence, which includes spillover effects or externalities, is “the existence of a functional relationship between what happens at one point in space and what happens elsewhere.”^65p11^ Spatial dependence seemed less likely to contaminate this analysis, but it was not implausible. For example, zero-sum dynamics could have emerged between neighboring units when vaccine doses were scarce. Similarly, peer-effects might have caused uneven diffusion of vaccination across space.

**Table e4.1:**
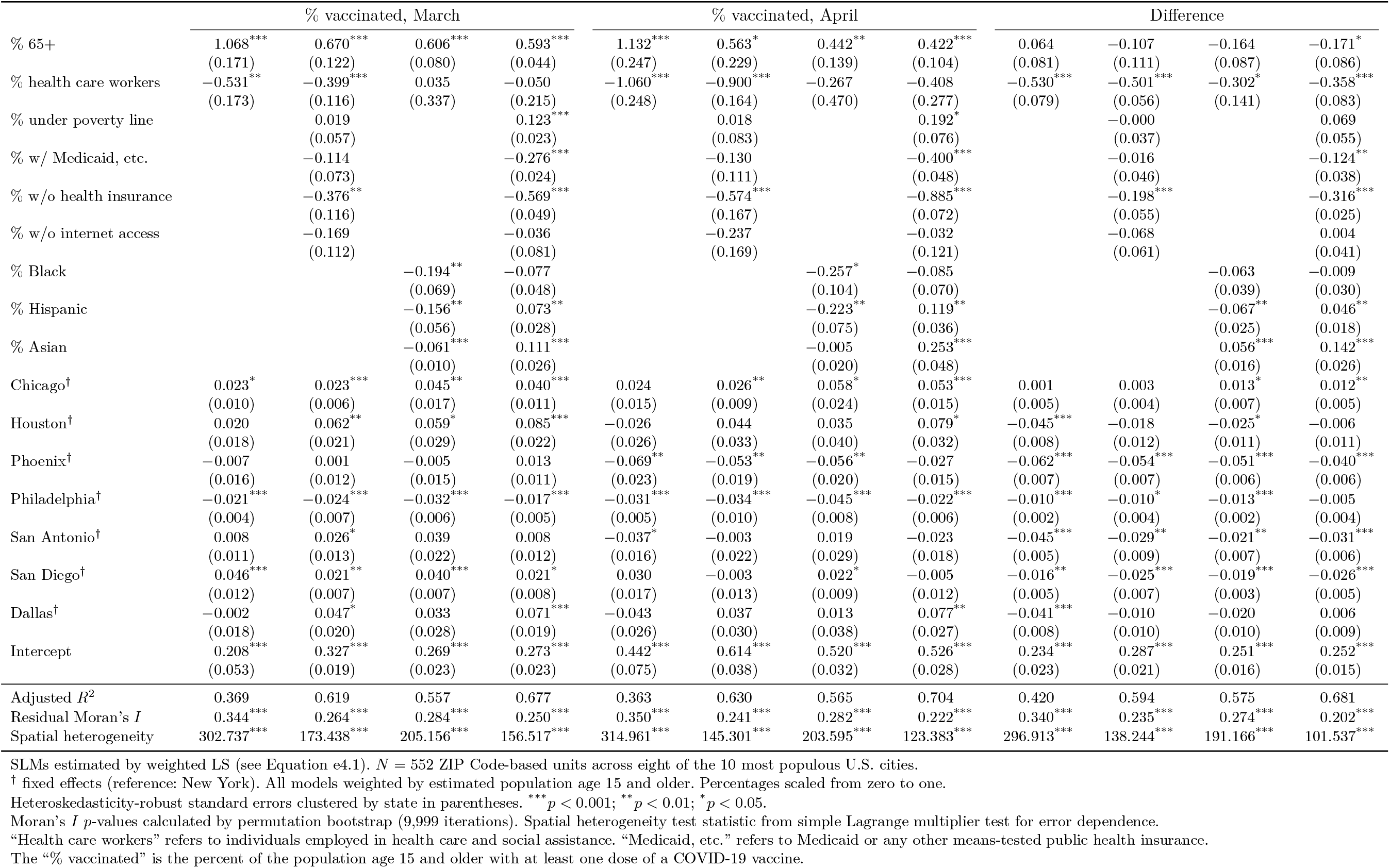
Step-wise standard linear model (SLM) estimates of COVID-19 vaccination levels in the population age 15 and older of ZIP Codes across eight large U.S. cities, March and April 2021

**Table e4.2:**
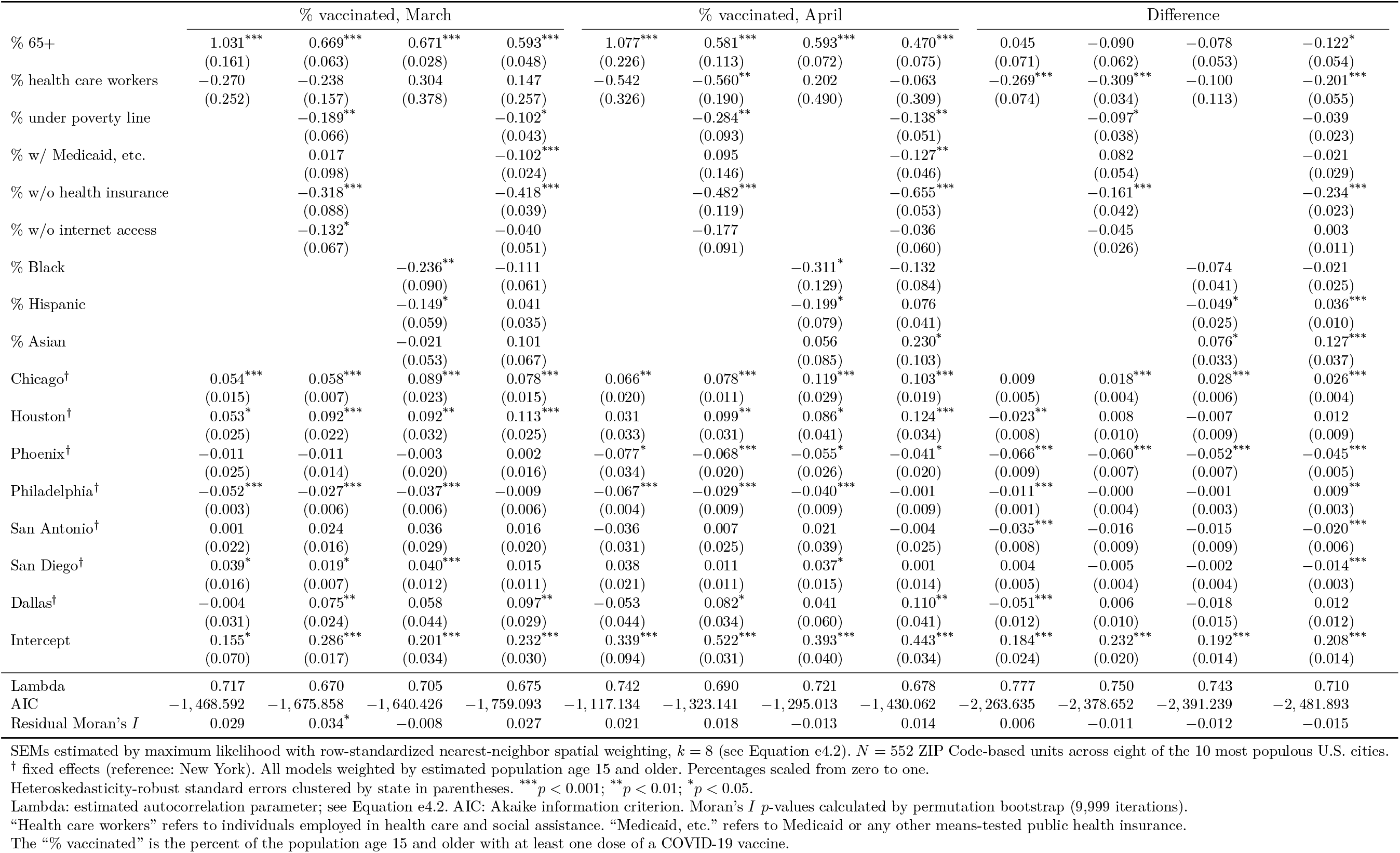
Step-wise spatial error model (SEM) estimates of COVID-19 vaccination levels in the population age 15 and older of interpolated units of analysis across eight large U.S. cities, March and April 2021

**Figure e4.1:**
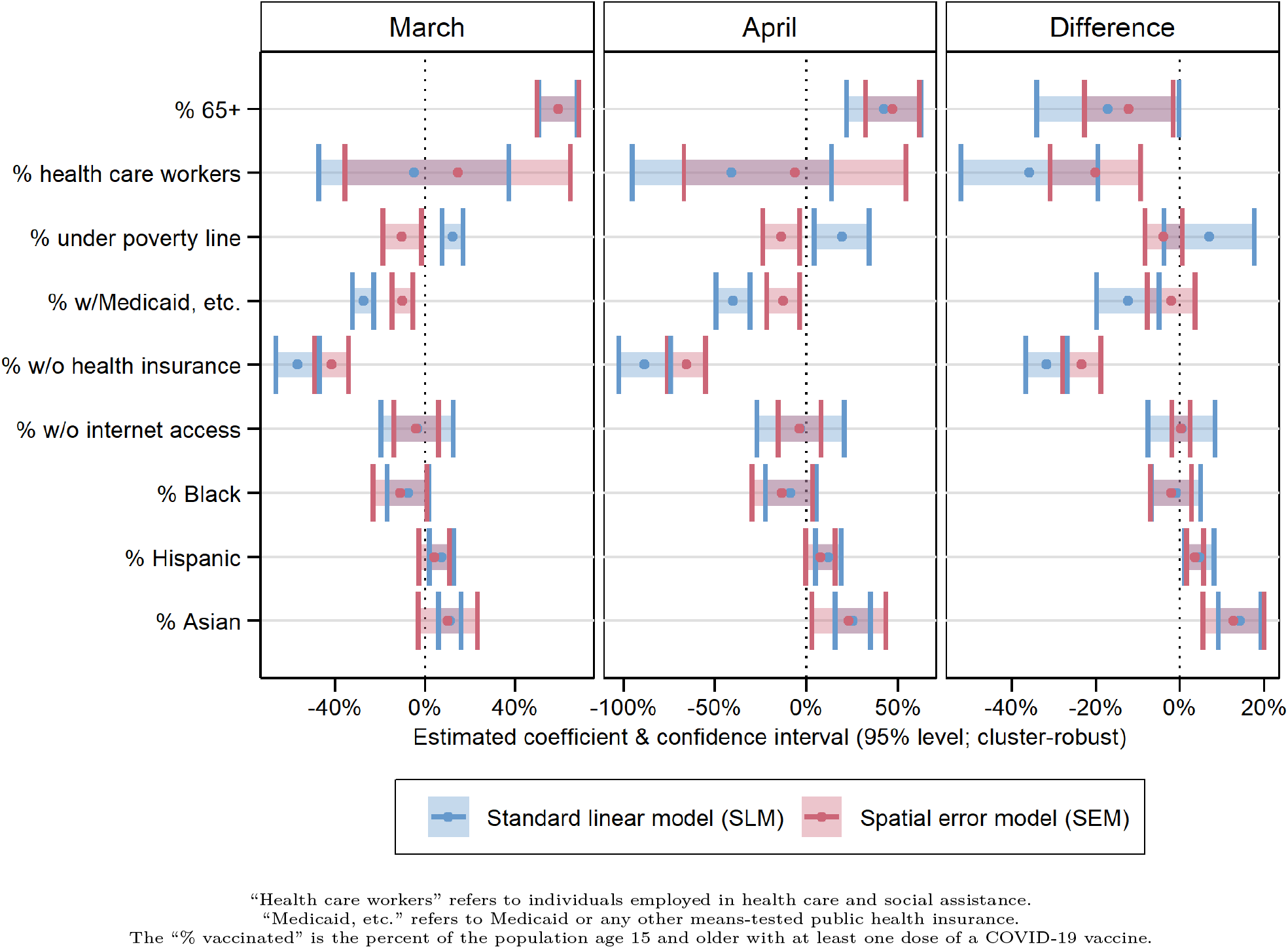
Estimated coefficients from standard linear model (SLM) and spatial error model (SEM) estimates of COVID-19 vaccination levels in the population age 15 and older of interpolated units of analysis across eight large U.S. cities, March and April 2021

Testing for and modeling spatial effects requires the researcher to specify the structure of spatial relationships. This structure is conventionally represented in matrix form as in Equation e4.4.^70–72^ We evaluated various common spatial weight matrix specifications, including adjacency (“queen” and “rook” contiguity), inverse distance, power distance, exponential distance, and nearest-neighbor weighting. We defined distance between units as the number of kilometers between their geometric centers, or centroids.^57, 73^

We ultimately used a row-standardized *k* nearest-neighbors scheme, with *k* = 8. With this specification of *W*, we assumed relationships were best measured relative to the eight other units closest to each unit. Each unit’s eight nearest-neighbors contributed equally to the spatial influence on it, and each unit’s spatial weights summed to one; each nearest-neighbor contributed one-eighth of the spatial influence on unit *i*.

In our multi-city analysis, nearest-neighbor weighting produced more consistent weights than contiguity- or distance-based weighting. The units of analysis were irregularly sized and sometimes physically discontinuous within cities, and density and sprawl varied considerably across cities (see Figure e3.1 and Table e3.4). Nearest-neighbor weighting ensured each unit was weighted relative to the same number of other units. It also avoided fluctuations from the varying spatial distributions of cities’ populations and the arbitrary boundaries of ZCTAs. Compared to contiguity-based weights, the nearest-neighbor approach was particularly advantageous for units on the edges of cities’ boundaries and units with few or zero adjacent neighbors (such as islands). Compared to distance-based weights, it avoided skew that could result from atypically large units that received artificially low weights due to their exaggerated inter-centroid distances with other units. Overall, nearest-neighbor weights with *k* = 8 struck an effective balance between the more rigid assumptions of contiguity-based weights and the perhaps overly-encompassing assumptions of distance-based weights.

We first tested for spatial effects by evaluating clustering in the SLM residuals using Moran’s *I*.^74^ A spatial complement to the conventional correlation coefficient, *I* measures spatial autocorrelation on a scale from negative one to one. To determine whether *I* was statistically distinguishable from zero, we ran permutation tests with 9, 999 iterations.^73^ We report Moran’s *I* for the SLMs that included all independent variables in the first row of Table e4.3. For each model, *I* was positive and highly significant, indicating units closer to one another had more similar residuals than units farther from one another (positive spatial autocorrelation). The Moran’s *I* test on the SLM residuals provided strong preliminary evidence of spatial effects, suggesting LS estimation was suboptimal in our setting.

Whereas *I* is a generic test statistic for spatial autocorrelation, Lagrange multiplier tests for SLM residuals help distinguish between types of spatial effects—i.e., to determine whether models exhibit spatial heterogeneity, dependence, or both.^75^ We summarize the results of the Lagrange multiplier tests for the SLM residuals in the bottom four rows of Table e4.3.^73^ The tests rejected the null hypothesis that the SLMs were free of spatial heterogeneity at *p <* 0.001. They retained the null hypothesis that the models were free of spatial dependence. These results held when testing for heterogeneity and dependence alone (simple tests) and when accounting for the simultaneous possibility of the other (robust tests). Lagrange multiplier tests strongly suggested the spatial relationships among our independent and dependent variables were heterogeneous across space, but no evidence emerged for dependence among nearby units.

**Table e4.3:** Tests for spatial effects in residuals of standard linear model (SLM) estimates of COVID-19 vaccination levels in the population age 15 and older of ZIP Codes across eight large U.S. cities, March and April 2021

| Test | March | April | Difference |
| --- | --- | --- | --- |
| Moran's $I$ | 0.250*** | 0.222*** | 0.202*** |
| Spatial heterogeneity (simple) | 156.517*** | 123.383*** | 101.537*** |
| Spatial dependence (simple) | 2.879 | 1.990 | 2.001 |
| Spatial heterogeneity (robust) | 153.982*** | 121.680*** | 99.808*** |
| Spatial dependence (robust) | 0.345 | 0.286 | 0.278 |

\*\*\* $p < 0.001$ ; \*\* $p < 0.01$ ; \* $p < 0.05$ .
Moran's $I$ $p$ -values from permutation bootstrap (9,999 iterations).
Simple Lagrange multiplier tests evaluate one spatial effect.
Robust Lagrange multiplier tests allow for simultaneous effects.

When spatial heterogeneity is present and spatial dependence absent, the SEM is a standard modeling approach.^65, 66, 72^ SEMs account for unobserved, spatially clustered independent variables—spatial autoccorelation in the disturbances—that lead to spatial clustering in the associations among observed variables. LS estimation cannot capture these spatial dynamics. SEMs yield reliable estimates in the absence of spillover effects and reduce bias from confounding variables.^69^ Another advantage of SEMs is interpretability. Unlike other spatial models, the independent variables are identical between SLMs and corresponding SEMs. The difference between SLMs and SEMs instead lies in the specification of the disturbances: SEMs introduce a spatially correlated vector of random effects, *u* in Equation e4.2. Interpreting SEM parameter estimates is thus comparable to interpreting SLM coefficients.^76^ By estimating SEMs, we assumed that *u* was independent of *X*—i.e., that unmeasured spatially clustered variables were uncorrelated with the observed independent variables.

##### City fixed effects, population weights, and cluster-robust standard errors

There were three other key components to our modeling strategy. First, we included city fixed effects. Because state and local authorities oversaw vaccine distribution and local political contexts varied dramatically, potential sources of city-level heterogeneity abounded. The fixed effects adjusted for unobserved characteristics that units shared within cities.^77^ They accounted for the possibility that average outcomes varied by city and purged associated confounding.

Second, we weighted the units of analysis by population using matrix *Q*. Population weighting addressed heteroskedasticity stemming from associations between the variance in vaccination outcomes and the number of residents of each unit.^78^ The substantive assumption of unweighted models would have been that each unit represented an equal share of the process of vaccine distribution and uptake. Estimating sample-wide average associations among the independent and dependent variables required weighting by the population of each unit that could receive the vaccine.^79–81^ Due to the weighting, more populous units and cities influenced estimation more than less populous counterparts, consistent with their greater share of the sampled population. Table e3.5 contextualizes the relative influence of each city.

Third, we assessed statistical significance using heteroskedasticity-robust standard errors clustered by state (cluster-robust standard errors). While fixed effects netted out city-level unobserved heterogeneity, disturbances for units in the same state were vulnerable to clustering. (Of the eight cities, three were in Texas). We addressed this issue by computing standard errors using the product of the conventional “sandwich” estimator^82^ and

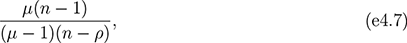

where *µ* = 6 was the number of clusters.^83, 84^ (This adjustment to the “sandwich” estimator is the default in many Stata commands). Calculated in this way, the standard errors were robust to within-state residual correlation.

#### e4.3 Simulations

##### Motivation

To illustrate relationships among the independent and dependent variables, we used a simulation approach. It was similar to conventional marginal effects analyses, which contextualize associations by visualizing the outcome that a model would predict given relevant values of the independent variables.^85, 86^ Because SEMs account for unobserved, spatially clustered variables, it is effectively impossible for researchers to identify appropriate points in the distribution at which to analyze trends and to predict the corresponding outcomes.^87, 88^ One approach would be to ignore the spatial structure of the disturbances and compute unit-level predictions for hypothetical observations, effectively treating the SEM as a SLM. This approach, however, distorts the modeled relationships.

Our simulations accounted for the spatial structure of the data. Instead of an estimate for individual hypothetical units as in marginal effects analyses, we derived sample-wide average estimated outcomes under instructive hypothetical conditions. We assumed every observation in the sample took on representative values at points of interest in the within-city distributions of the independent variables. We could thereby answer a simple counterfactual question: if every unit sat at the same place in its city’s population distribution and were subject to the trends suggested by the SEM estimation, how would average vaccination outcomes compare to the observed averages? Figure e4.2 illustrates expanded results of the simulations.

As we indicate in Section e2.2, this simulation approach contextualized disparities more comprehensively than interpreting coefficients alone. Table e4.2 and Figure e4.1 show that the outcomes’ estimated associations with percents Black and Hispanic were statistically insignificant in March and April; average vaccination outcomes did not change systematically with Black and Hispanic populations, conditional on vaccine priority populations, socioeconomic composition, and the spatial relationships specified in *W* . In isolation, these results suggest that economimc—not racial/ethnic—inequality explained variation in vaccination outcomes. By accounting for unequal distributions of SES and vaccine priority populations across racial/ethnic groups, however, the simulations revealed that the explanation is more complicated. Because economic inequality is racialized, areas with high Black and Hispanic populations lagged behind areas with high Asian and White populations—even though the coefficients for percents Black and Hispanic were insignificant and the latter was even slightly positive. Attending to direct and indirect channels of structural racism more accurately represented the inequality-generating process than focusing only on average conditional racial/ethnic disparities.

**Figure e4.2:**
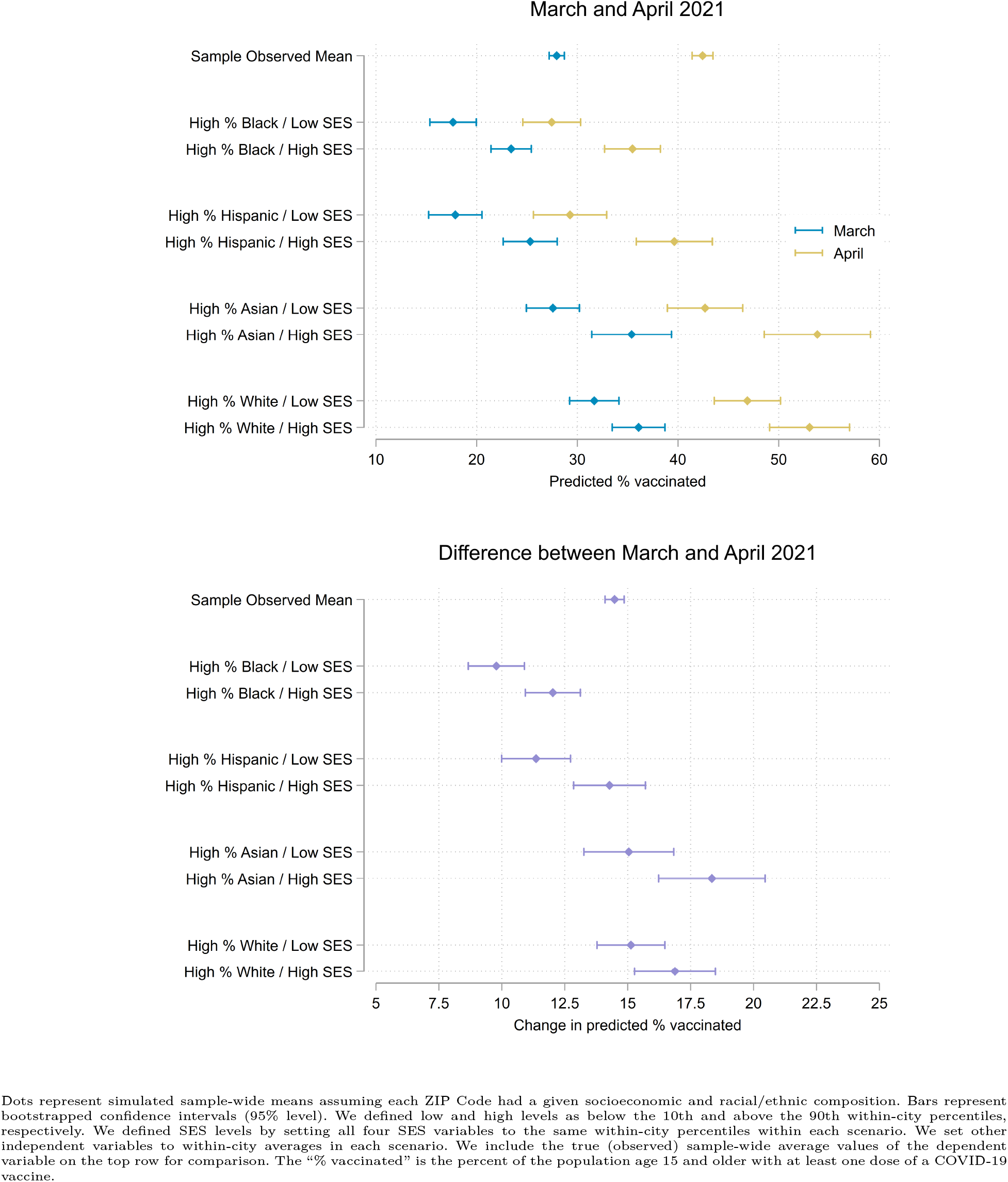
Simulated COVID-19 vaccination levels by racial/ethnic and socioeconomic composition in the population age 15 and older of ZIP Codes across eight large U.S. cities, March and April 2021 (expanded)

**Table e4.4:**
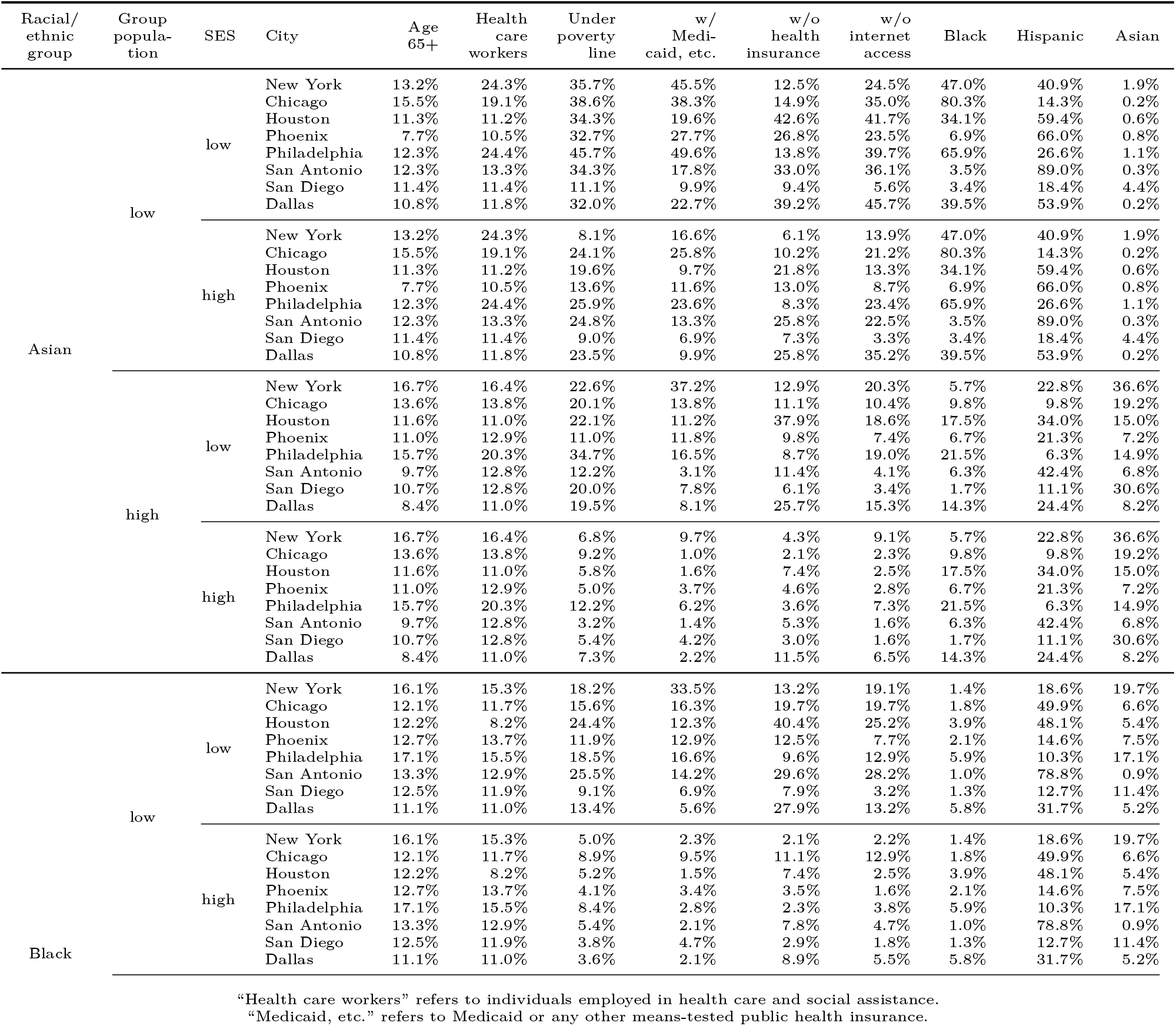
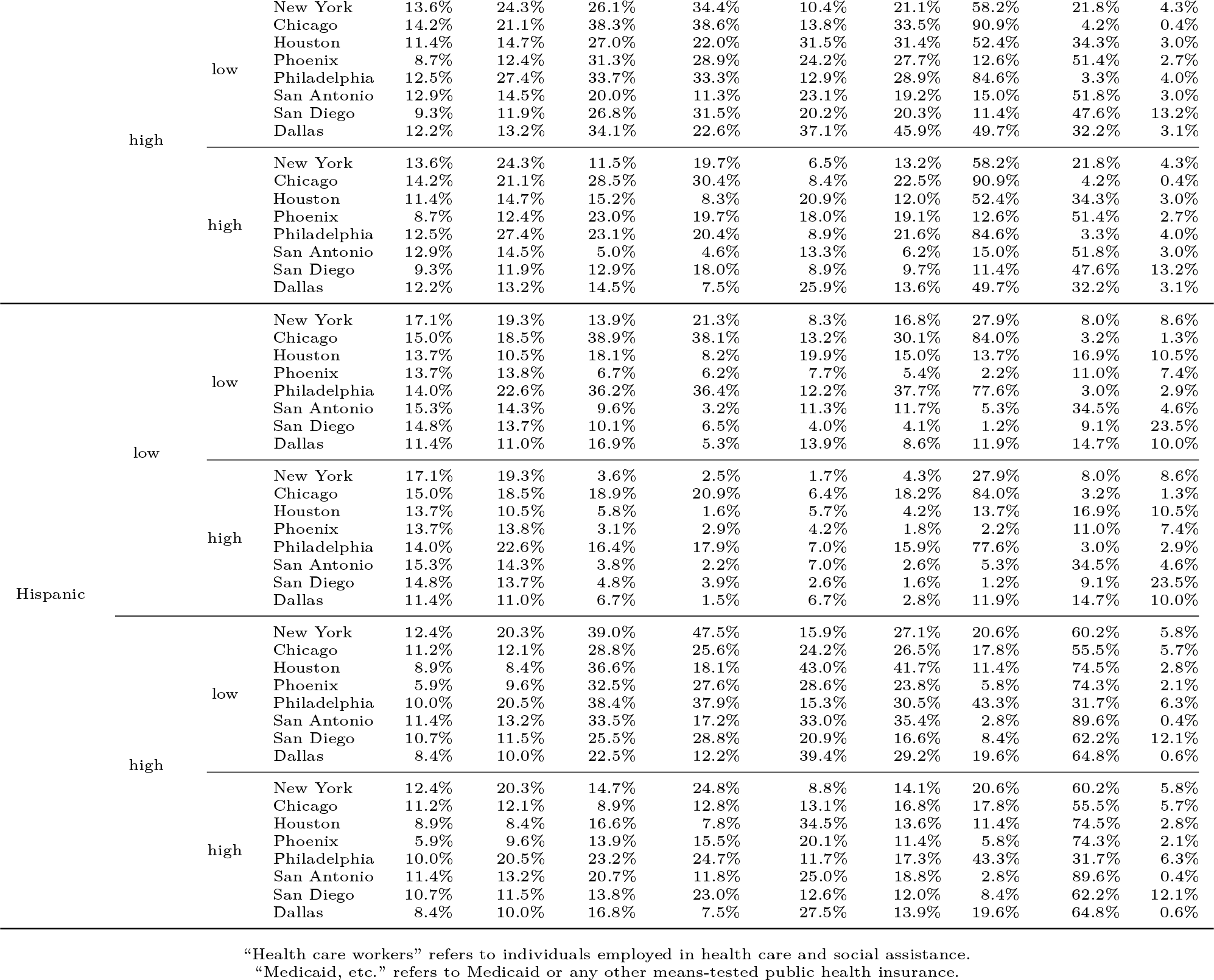
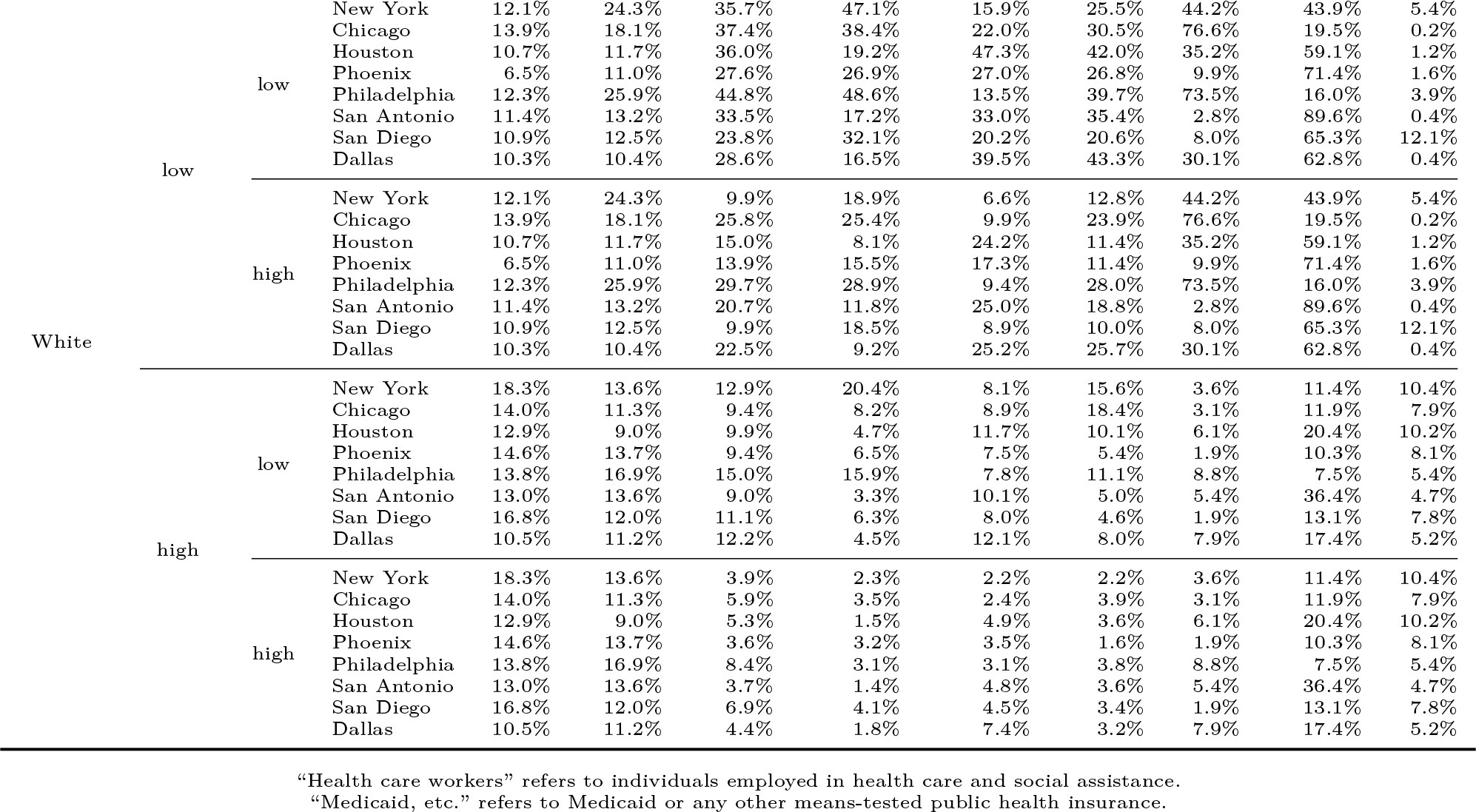
Plausible values for independent variables used in simulations of COVID-19 vaccination levels by racial/ethnic and socioeconomic composition in the population age 15 and older of ZIP Codes across eight large U.S. cities, March and April 2021

##### Implementation

We first identified plausible values of the independent variables, listed in Table e4.4. Within each city, we identified low and high levels (the 10th and 90th percentiles) of Black, Hispanic, Asian, and White populations. Given these racial/ethnic compositions, we set plausible low and high values of the four SES variables. We identified the within-city 10th and 90th percentiles of the SES variables among units with populations within five percentile points of the within-city low and high levels for the given racial/ethnic group (the fifth through 15th and 85th through 95th percentiles). We set all other independent variables to their weighted averages within the same ranges. For example, in the iteration identified in Figure e4.2 as “High % Black / Low SES,” we assigned all units low values (10th percentile) for the SES variables and average values of other independent variables, among units with high Black populations (85th through 95th percentiles) within their respective cities.

Next, we simulated outcomes given the plausible values of the independent variables. For each combination of racial/ethnic and socioeconomic compositions, we set the variables for each unit in our data set to the corresponding within-city values in Table e4.4. (We assigned every unit within each city the same values for each independent variable in each iteration). For each unit, we then calculated the estimated values of the outcome under each scenario, using the parameter estimates from the full models in Table e4.2. We computed population-weighted means for the three outcomes under each hypothetical scenario.

Finally, we bootstrapped confidence intervals for the simulated sample means. We obtained 1,000 resamples of our original data set by sampling it with replacement.^89^ We then adjusted the spatial weights matrix *W* accordingly for each resample. We assigned nearest neighbors according to *h_i_* in the original sample, which maintained the spatial structure of the data across iterations. Next, we re-estimated the SEMs on each of the resamples. We simulated the outcome variables under each hypothetical population composition of interest by assigning each unit the corresponding value in Table e4.4 in the same manner as above. From the distribution of the 1,000 resulting population-weighted resample means for each outcome, we calculated pivot confidence intervals at the 95% level.^90^ This bootstrap procedure yielded a non-parametric approximation of the uncertainty in the hypothetical outcomes.

## Footnotes

e We detail how we interpolated units of analysis that could be meaningfully compared given agencies’ diverging reporting practices in Section e3.2.

f We further introduce ZCTAs in Section e3.1.

g We introduce CBGs in Section e3.2.

h We introduce ZCTAs in Section e3.1.

i As the third column of Table e2.2 shows, the ACS tables were sampled from different sub-populations. Variation in the universes was slight and unproblematic for our analysis.

